# A Genome-Wide Association Study (GWAS) meta-analysis of post-traumatic osteoarthritis of the knee (GO-PTOA): Protocol for Analysis Plan

**DOI:** 10.1101/2023.10.20.23297162

**Authors:** Merry-Lynn McDonald, Joshua Richman, Cindy Boer, Raneem Kalsoum, Daniel Dochtermann, Tyler Barker, Kristin Briem, Konstantinos Hatzikotoulas, Benjamin Hollis, Luke Jostins-Dean, Reedik Magi, Ene Reimann, EstBB Research Team, Lorraine Southam, Unnur Styrkarsdottir, Hemant Tiwari, Joyce Van Meurs, Eleftheria Zeggini, Jasvinder Singh, Fiona E. Watt

**Affiliations:** University of Alabama at Birmingham, Birmingham, USA; Erasmus University, Rotterdam, Netherlands; Centre for Inflammatory Disease, Department of Immunology & Inflammation, Imperial College London, London, UK; Wexner Medical Center, The Ohio State University, Columbus, Ohio, USA; Department of Orthopaedics, University of Utah, Salt Lake City, Utah, USA; Department of physical therapy, University of Iceland, Iceland; Institute of Translational Genomics, Helmholtz, Munich, Germany; Centre for Osteoarthritis Pathogenesis Versus Arthritis, Kennedy Institute of Rheumatology, University of Oxford, UK; Institute of Genomics, University of Tartu, Estonia; DeCODE Genetics, Iceland

**Keywords:** Osteoarthritis, Post-traumatic osteoarthritis, Genome Wide Analysis Study

## Abstract

Post Traumatic Osteoarthritis (PTOA) of the knee is osteoarthritis (OA) occurring specifically after a significant acute injury to the joint. Approximately 50% of people with significant knee joint injuries, such as anterior cruciate ligament (ACL) rupture, develop symptomatic radiographic OA within 10 years. Although impacts of the disease are well-described, there is little known about the genetic risk of developing OA after a knee injury relative to idiopathic OA. There is an unmet clinical need to understand the aetiology of PTOA, which will help us to understand, predict and prevent PTOA.

In this Analysis Plan, we describe our methodological approach to performing a Genome Wide Association Study (GWAS) meta-analysis, developed by the Genetics of Osteoarthritis consortium (GO)-PTOA working group. We aim to determine if genetic variants associate with PTOA following a knee joint injury and understand whether these variants are similar to or different from those associated with idiopathic knee OA (iOA).

Summary statistics from six international cohorts and biobanks will be included in the meta-analysis. We have harmonized an approach to identify cases of acute knee injury, knee PTOA cases as well as controls. Our three main objectives are to identify genetic variation associated with:

1. knee PTOA independent of knee iOA
2. knee PTOA compared with knee injured controls without OA
3. knee PTOA compared with controls over 40 years of age without injury or OA Although not part of our primary analysis, we also plan to identify genetic variation associated with the onset of knee PTOA in a time-to-event analysis in the future.

Where resources allow, sensitivity analyses for all aims will be carried out, adjusting for body mass index (BMI). For case-control analyses, either a logistic regression or linear mixed model (LMM) will be used. For later time-to-event analysis, a matched case-control Cox Proportion Hazards (PH) regression model will be applied. Cohorts will share their summary statistics with the central analysis site conducting the single meta-analysis for each GWAS, which is likely to be carried out in either METAL or GWAMA.

Findings from this work will potentially improve our understanding of PTOA pathogenesis and aetiology. Better understanding of genetic determinants could contribute to more accurate prognostic modelling and precision medicine in the future.

## 1.0 Objective of GO-PTOA

The objective of GO-PTOA is to identify genome-wide significant signals associated with post-traumatic osteoarthritis (PTOA) of the knee.

https://www.genetics-osteoarthritis.com/active-working-groups/gwas-meta-analysis-of-post-traumatic-osteoarthritis-of-the-knee/index.html

## 2.0 Purpose of this document

This plan has been written to reflect our approach to our planned work, supported by meetings of the GO-PTOA working group. In this document we cover the following:

- Background to the work we will carry out and its relevance
- Our main research questions and objectives
- The overarching principles in our methodological approach, including methods for case ascertainment and eligibility criteria
- The statistical approach and meta-analysis
- Publication plans
- Management of results including publicly available deposit of summary statistics and scripts post-publication

## 3.0 Background to work

Post-traumatic osteoarthritis (PTOA) of the knee is the osteoarthritis (OA) that follows clinically significant acute knee injury in a proportion of people. It is thought to affect around 50% of people experiencing a knee injury, such as anterior cruciate ligament (ACL) injury or acute traumatic meniscal tear. It comprises about 12% of all people with knee OA, but causes particular impact as it affects people earlier in their life course. The polygenic risks of knee OA as a whole are now well known. However, very little is understood about the genetic risk of developing OA after a knee injury. This is important for our understanding of OA as a whole and for translational research in OA (where pre-clinical modelling relies heavily on post-traumatic models, assuming that this disease is one and the same).

In summary, there is an unmet clinical need to understand, predict and prevent PTOA. A better understanding of the aetiology of PTOA, including genetic risk of PTOA is likely to give insight into its pathogenesis and that of OA. Understanding genetic determinants may contribute to more accurate prognostic modelling and to precision medicine in this area. Beyond this, determining the similarities and differences between non-traumatic ‘idiopathic’ OA (iOA) and PTOA are vital for the field as a whole, particularly for successful translational research and progress towards better tests and treatments for people living with or at risk of this condition.

## 4.0 Research questions

Our main question is to determine whether there is genetic variation associated with the development of PTOA after a knee joint injury.

**Our overall hypothesis** is that there is genetic variation associated with the development of PTOA after a knee joint injury (PTOA). We want to definitively answer whether these variants are the same as, or different from idiopathic knee OA (iOA) genetic variants.

**Our aims** are to carry out several linked GWAS meta-analyses to answer this question.

We will aim to show whether the genetic architecture of PTOA is the same as, or different to iOA.

## 5.0 Overarching principles in methodological approach

Our aim is to include as many GO cohorts/biobanks/collections (henceforth referred to as ‘cohorts’) to participate in the analysis as possible, in order to increase numbers of international cohorts and participants, and therefore our power and generalisability in answering our research questions.

We will do this by accepting some loss of precision in case ascertainment and allow increased heterogeneity in our methods of defining cases, whilst using a prior approach in UK Biobank as a starting guide as to how we might conduct a meta-analysis at scale.

Given the high frequency and impact of PTOA of the knee, that these cases can be identified from electronic data and also to increase homogeneity in analyses, our study will be exclusively in the knee.

The analysis will be carried out in two stages. Firstly, the consortium will work with each participating cohort to develop an approach to identify PTOA cases and appropriate controls so that each cohort can run their own individual GWAS studies (pre-defined case-control, and where possible also time-to-event) according to the principles laid out in this document. Secondly, each cohort will share the genome-wide summary statistics for their analyses with a single coordinating ‘central analysis site’, who will then carry out a single meta-analysis for each GWAS according to a pre-defined plan.

We will aim for **definition of PTOA knee cases** in one or both of the following ways:

1. Evidence of at least one clinically significant knee injury in a participant*, which is likely due to acute trauma to an internal joint structure (at/within the knee joint) rather than elsewhere AND a subsequent OA code which is likely of the same knee (or of either knee if side is not known)
2. Evidence of a specific diagnosis of PTOA of the knee

*When considering evidence for a clinically significant knee injury:

- This can be a range of knee injuries, but ideally without evidence of more extensive trauma (see Eligibility criteria, Appendix 1 & 2)
- A recall of a ‘history of knee injury’ by a patient that is not specific to a particular clinically assessed structural knee injury is not sufficient, without additional supporting evidence
- A clinician diagnosed injury of a relevant structure, supported by appropriate imaging evidence of an injury, and/or supported by a single diagnostic code or combination of diagnostic codes, such as on ICD-10 is sufficient

### Participation in GO-PTOA requires cohorts

- To be able to identify group i) listed below AND at least one of groups ii-iv), but ideally as many groups as possible
- To have the ability to undertake these analyses* and supply the summary statistics by the agreed date in 2023

*typically this is anticipated to be local analysis, or with support from other participants’ teams e.g. Masters students within GO-PTOA, where this has been arranged in advance

The following groups shown in Figure 1 are mutually exclusive for the purpose of GWAS case-control analyses:

i. Post-traumatic knee OA cases (as defined above) ‘<u>PTOA’</u>
ii. Non traumatic/’idiopathic’ knee OA (knee OA with <u>no</u> evidence of prior knee injury, however this is defined) ‘<u>iOA’</u>
iii. Knee injured, no knee OA (after as much follow up as is possible) ‘<u>Knee injured controls’</u>
iv. Injury free, OA free controls (age/sex matched where possible to cases of injury for the relevant analyses) ‘<u>Injury free, OA free controls’</u>

**Figure 1:**
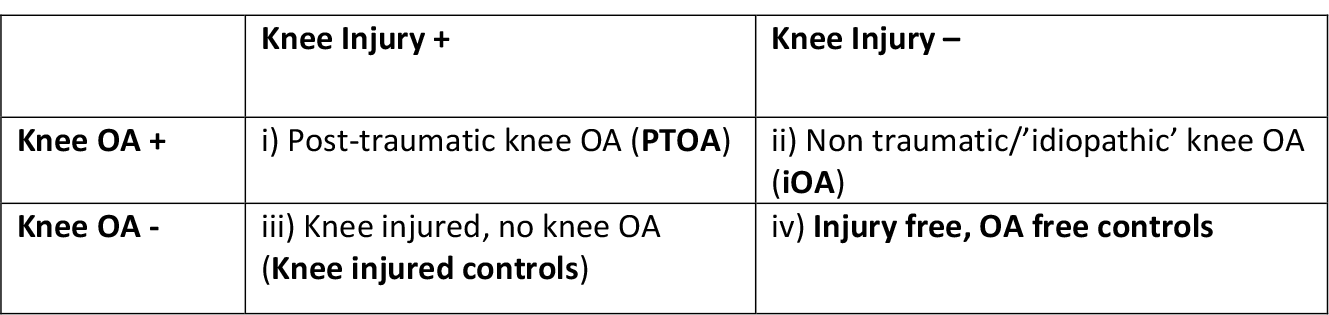
GWAS case control analysis groups. A summary ‘knee injured group’, which is the combination of i) and iii) is relevant for some analyses: these are all those who have been knee injured, irrespective of their OA outcome ‘ALL Knee injured’.<u>For inclusions and exclusions for each group/analysis, see Appendix 5.</u>

### 5.1. Case ascertainment and eligibility criteria: guiding principles

Methods for case ascertainment should be broadly informed by/based on prior work in UK Biobank (UKB) by some of this group and the discussions of the GO-PTOA working group at their first three meetings (see eligibility criteria and code lists, Appendix 1 and 2, meeting minutes).

Each cohort should consider the inclusion/exclusion criteria below and carefully document their approach and inclusion strategy, for example, by describing what types of injury are likely to have been included. However, the general guidance will be to allow different approaches and that, where these are felt likely to potentially represent substantially different groups, sensitivity analyses will be carried out. For example, if some inclusion approaches are more likely to be associated with associated fractures/more extensive trauma, analyses could capture susceptibility to bone fracture etc.

Where core/shared Eligibility criteria by participating cohorts can be retrospectively defined, these will be captured, or else summarised per cohort at reporting.

#### 5.1.1. Inclusions: Knee injured

Considering the above and Inclusion Criteria from UKB analyses (Appendix 1A), a simplified/subset of inclusion criteria for definition of acute knee injury and for subsequent/related definition of PTOA of the knee will be outlined by each cohort.

Inclusion criteria will be considered, adapted and code lists or definitions refined and reported by each cohort for each of the 2-4 groups for this purpose (we will report at cohort level, as approaches will necessarily vary).

Cross-mapping (new or existing) from ICD-10 to other coding systems such as ICD-9 may be carried out to support this work. The need for this should be identified as soon as possible.

#### 5.1.2. Exclusions (all groups)

Considering the Exclusion criteria for the UKB study (Appendix 2), adaptation of these can be defined by the cohort.

Where possible for the time-to-event analysis, these should be relative to the injury event, as described in Appendix 2A.

As a minimum, for all analyses, these should include as a minimum the ‘ever’ exclusions in this list of inflammatory arthritis, multiple site trauma, congenital bone disease, etc. as listed in Appendix 2B (these are a subgroup of Appendix 2A, items 6-12).

##### These exclusion criteria should be applied to all 4 groups

For exclusions for group (i) PTOA cases only, where the presence of both injury codes and OA codes are used to define PTOA cases, it is important that in addition those with knee OA occurring <u>before</u> the injury, i.e. exclusion criterion 1, are excluded.

#### 5.1.3. Knee OA outcomes

Knee OA codes: Codes/diagnoses compatible with ‘definite, probable or possible knee OA or total knee replacement/revision consistent with knee OA’ including codes consistent with post-traumatic OA knee (see Appendix 3A).

- Exclusions are if OA is of lower limb but not of the knee e.g. hip, ankle, or OA which is definitely not of lower limb. (Knee, hip and ankle location codes can be used where relevant, to improve specificity).
- Timing of OA outcome (where relevant) should be the earliest documented knee OA code.
- OA diagnosis should be <u>6 months or more</u> after the knee injury.

### 5.2. Time dependent vs time independent analyses

Cohorts will fall in to two main groups in terms of case ascertainment for group (i) ‘PTOA’ in terms of whether they can define the exposure, injury and the outcome OA separately.

Participating cohorts need to be clear which they belong to (it is also possible for a cohort to contribute both types of cases to the analyses, in which case the two groups should be mutually exclusive):

*Where the knee injury is defined/timed and followed with a subsequent timed OA code/diagnosis or lack of OA code. In these cases:*

- The knee injury (defined as supported by Appendix 1A) should be between the ages of **16** (or 18 if lack of consent for this) **and 50 inclusive**
- An upper limit for the observation period of <u>20 years after the knee injury</u> should be applied for cases of subsequent OA, or censoring to that date (or at death or last record if sooner)
- OA outcomes can include PTOA knee codes (see 5.1.4)
- Pre-existing OA prior to the injury or conditions which suggest this should be excluded (as supported by Appendix 2A)
- Knee OA diagnosis in the 6 months after injury ascertainment should be excluded (as likely pre-existing OA, see 5.1.4)
- These cohorts can contribute time-specific content to a time-to-event analysis if they are able to, as well as identifying so-called ‘time to event’ cases to contribute to the case-control analyses.

*Where there is no time stamp/specific record of the knee injury:*

The main example here is where there are combined ‘PTOA knee code cases’ (for example, as found in Appendix 1B) where there is no available prior information on what the knee injury was. In these circumstances, inclusions and exclusions may be simplified by the cohort (as cannot be relative to the timing of an injury).

Specifically:

- Age at time of injury may not be considered (any age at code is acceptable)
- Time between injury and OA cannot be considered (just clinical judgement that this is PTOA)
- Non time dependent exclusions (Appendix 2B) should still be applied

*Where cohorts are providing both types of cases from the same cohort:*

Only PTOA knee code cases that have <u>not</u> been identified in the time to event analyses should be supplied i.e., these two groups should be mutually exclusive.

Where a cohort has both methods of identifying PTOA cases, for primary analyses 2 and 3 (section 5.4), ‘time to event’ methods should be prioritised and only <u>additional cases</u> from PTOA knee codes not already identified should be <u>combined with those identified in time-to-event analyses.</u>

### 5.3. Other variables and baseline characteristic reporting

Age at time of injury and sex are relevant covariates in the time-to-event analysis (i.e. they influence the outcome, the development of PTOA). As such, wherever possible, they should be used as covariates in time-to event analyses. Covariates therefore include:

- Age at time of injury
- Or, Age at time of inclusion (diagnosis/last follow up – see analysis section)
- Sex
- Global ancestry group (if different ancestral groups are included)
- First X genetic principal components (as defined by cohort)
- Any other cohort specific covariates, felt to be important by cohort, such as Genotyping chip

For all analyses, the following summary information should be supplied for the purposes of reporting on baseline characteristics (Table 1) for each cohort and for each study in the meta-analysis:

- Current age (median, IQR)
- Age at time of injury if known (median, IQR)
- Sex distribution (n/% female)
- BMI (at time of injury or other time point (report when in Appendix 5 tables) (median, IQR)
- Global ancestry group (if different ancestral groups are included)

### 5.4. Power considerations and sample sizes

Power calculations using UKB data indicated that for a full meta-analysis, a total sample size between 6,000 and 9,500 injury cases would be required to detect variants in a time to event GWAS with moderate hazard ratios (HR 1.2-1.25; assumed MAF = 0.3) and with power of 80%, at genome-wide significance.

To compare PTOA and iOA associations in a full GWAS meta-analysis, odds ratios of >1.2 could be detected at genome-wide significance with sample size of 2,750 PTOA cases (assuming an excess of iOA cases), though greater case numbers are preferable as they allow detection of smaller effect sizes.

Based on completed feasibility assessments within GO-PTOA participating cohorts (April 2022-February 2023), it is very likely that these sample numbers will be exceeded.

There will be no minimum sample size of PTOA cases in a participating cohort/collection to take part (though some cases will be necessary). However, those with smaller sample sizes may be combined (where genotypic data can be shared/accessible – noting this would not be typical or anticipated for the majority) or may be subject to sensitivity analysis, given that their inclusion may introduce a disproportionate amount of heterogeneity.

### 5.5. Analysis approach

One or more of these defined GWAS will be performed by the participating cohorts themselves, depending on their dataset and ability to carry out these analyses, prioritising as listed below. Meta-analysis of summary statistics from all contributing cohorts for that particular analysis will then be carried out by the Central Analysis Site for GO-PTOA.

The group’s Analysis Lead will oversee the delivery of the meta-analysis at the Central Analysis Site. Regular meetings will be arranged to support those contributing data to the analysis with project management by the coordinating Imperial College team. A practical document for the primary analysis is included as Appendix 5, in support of this plan.

#### Primary GWAS models*

The following analyses have been prioritised by the group, in this order. See Appendix 5 for inclusion/exclusion criteria for all case and control groups.

- A case-control GWAS of all PTOA cases vs. iOA
- A case-control GWAS of all PTOA cases vs. knee injured controls (no OA) <u>without</u> considering time to event/censor
- A time-to-event GWAS of all knee injured cases with available time information, looking at the subsequent event of presence of OA/PTOA, to 20 years of follow up or censor)*

#### Secondary GWAS models

There will be 3 additional analyses, where cohorts have capacity:

- A case-control GWAS of PTOA vs. Injury free, OA free controls
- A case-control GWAS ALL Knee injured, no OA vs Injury free, OA free controls (to interpret any findings in (4) this will be necessary)
- A case-control GWAS of iOA vs. Injury free, OA free controls (to support post GWAS analyses)

*to allow as many participants as possible to take part in the first primary analyses and maximise our power and achievability of our first analyses, <u>case-control GWAS by participating cohorts (first, then second, fourth and fifth in the order listed here) have been prioritised for sequential analysis and submission,</u> over the time to event analysis. The time to event analysis will be performed after these are case control analyses are complete, by those cohorts able to do this, informed by these first analyses.

Post GWAS analyses will be carried out in certain cohorts (nominated after combined meta-analysis, informed by the results from primary and secondary GWAS analyses above. A separate detailed post GWAS analysis plan will be written which will support these specific analyses. These are linked analyses and it is intended that these will include:

- **The predictive effects of polygenic risk scores (PRS) for iOA** knee or total (any joint) OA **on PTOA** will be assessed by creating and adding them as covariates in the time-to-event analysis in 1. (Exact methods to be confirmed but wherever possible, there will be cross-replication e.g. using iOA scores from one cohort in PTOA analyses of another cohort, to ensure generalizability of the scores.) This is likely not to be in the largest or smallest sample populations and would aim to include PRS from non-White populations.
- **Testing the coheritability between PTOA and iOA of the knee**. We will seek to quantify the shared and non-shared genetic risk for iOA and PTOA. Variance will be partitioned and quantified into that shared between PTOA and iOA, and that which is not co-heritable, by genome-wide LD score regression analysis applied to genome-wide summary statistics for PTOA knee (log hazard ratios or log odds-ratios) and iOA knee (log odds-ratios). We will test for heterogeneity, finding out if any effects seen are greater/lesser in PTOA.

### 5.6. Statistical approach

Our approach will use two main methods:

- For the time-to-event analysis, a matched case-control Cox Proportional Hazards (PH) regression model will be applied, implemented in SPACox (a module in R), GATE or similar. Scripts and methods will be shared with cohorts where this is being applied.
- For case-control analyses, either logistic regression or LMM will be used, at the discretion of the cohort, depending on what is most appropriate considering their population structure and size. Their selected method, software used and imputation methods should be reported by the cohort on the relevant tab on the Google sheet.

As per 5.3, covariates for both analyses will include genetic principal components (PCs), genotyping batch (where relevant), age at the time of injury (where available), sex and any cohort specific covariates (cohorts may determine the number of genetic PCs needed, with the aim of explaining approximately 80% of the variance as a benchmark, to make this as comparable as possible and report the numbers of these. The minor allele frequency will be >= 0.01).

Where used as an additional covariate in the post GWAS analyses, PRS for iOA (knee, any site) will be generated in cases that are independent to the time-to-event analysis.

### 5.7. Analysis by each participating cohort

Chip and method of imputation, number of genetic PCs, prior sample and SNP QC approach as well as inclusion/exclusion approaches will need to be summarised by each cohort.

Summary statistics for each GWAS carried out should include total sample size, chromosome, SNP position, HWE, allele1, allele2 (noting which is minor allele), frequency of allele 1 in cases, frequency of allele 1 in controls, beta/odds ratios (either log odds ratios/log hazard ratios relative to allele 1, depending on analysis), standard error (95% CIs), *P*-value (an example output .csv file will be provided to support their upload).

It is anticipated that PLINK will be used for GWAS analyses. R will be used for all other analyses. The output files may differ and file type should be notified to the coordinating site.

### 5.8. Meta-analysis approach

In all cases, the summary statistics from each of up to 6 GWAS analyses listed in 5.4 will be shared with the Central Analysis Site. The analysis team will use a fixed-effect variance weighted meta-analysis for each of the 6 analyses to produce 6 sets of meta-analysed summary statistics. (We will assume the same allelic effects over all studies but will test for heterogeneity between effects).

Analysis is likely to be carried out in METAL or GWAMA (this will be reviewed by the central analysis team, depending on the datasets), to enable checking for strand alignment.

GWAMA may be preferable for additional trans-ethnic meta-analysis. Forest plots and relevant tests (I2) will be used to assess for heterogeneity.

Genome-wide significance threshold will be corrected for multiple testing.

### 5.9. Sensitivity analyses

To check if the methods for case ascertainment influence the results, we will report effect sizes across different types of methods of identification and carry out predefined sensitivity analyses.

These will include one or more of:

- omitting sample size outliers, i.e. very large or very small cohorts, to check how these affect results
- omitting certain ancestry groups (or if sample numbers for these subgroups allow, running specific meta-analyses within these)
- only considering variants found in at least 2 cohorts with the same direction of effect
- sensitivity analyses including BMI as a covariate
- sensitivity analyses excluding individuals/cohorts where there is a) a PTOA case with no prior knee injury information, i.e. PTOA case codes only and b) insufficient follow up of knee injured comparators

For details on sensitivity analyses, please see Appendix 5.

## 6.0. Post-meta-analysis activities

Fine mapping will be carried out for each independent signal, including all variants within 1Mb of the index variant, to extract potentially associated variants. Manhattan plots and Locus Zoom plots will be prepared. Approaches such as SuSiE will be used to generate a credible set. Ancestry specific and trans-ethnic fine mapping may be used, depending on the constituent cohorts.

We will tabulate hits with genome wide significance and search across open access databases including Open Targets Genetics Portal to identify candidate genes and other phenotypes associated with variants, reporting our findings.

We will aim to carry out independent replication of any findings in other replication cohorts not taking part in the GO-PTOA primary analysis.

## 7.0. Access, review and storage of results

### 7.1. During the GO-PTOA primary analysis

Summary statistics from each cohort in the agreed output format will be uploaded to a user-restricted Box account, supported by the Central Analysis Site. Genome-wide summary statistics for each meta-analysis generated during this study will be held in a central repository at the Central Analysis Site during the analysis. This will be a password protected secure private Github, with vetted access only to those who are members of the GO consortium (who have signed or agreed to abide by the principles of the GO memorandum of understanding) and who are approved by the GO consortium PTOA sub-project leaders (FW, analysis group lead). Summary statistics for cohorts will be made accessible to these approved members of the group to allow their combined analysis at consortium level, with transfer via SFTP between analysis servers where necessary.

The analysis and its results will be reviewed by the GO-PTOA group at each group meeting.

### 7.2. After publication of GO-PTOA primary analysis

Following completion of analyses, summary statistics will be uploaded to a repository, at the time of publication.

Individual summary statistics will be uploaded locally as per cohort guidelines/requirements. Following publication, they will be uploaded to MSK Knowledge Portal as per GO consortium procedures, to allow open access https://msk.hugeamp.org.

Final summary statistics for the meta-analyses will also be uploaded to the GWAS Catalog https://www.ebi.ac.uk/gwas/.

Summary statistics dropping out cohorts will be reviewed and may be necessary for upload for some local repositories.

It is intended that the analysis approaches, software/scripts and summary statistics will all be archived and publicly accessible when agreed by all group members, by public Github page, at the time of/following publication.

## 8.0. Publication

The results of this analysis will be published in a peer reviewed open access journal. Publication/authorship policy will be in line with GO collaboration agreement and ICJME guidance, agreed by the group and represent the contributions of the cohorts, analysts, as well as analysis lead and group lead. Near final drafts will be circulated to the GO steering committee and other scientific committees of relevant cohorts at least 3 weeks before manuscript submission.

## 9.0. Coordination

Please see the consortium’s memorandum of understanding (January 2023) for guidance on consortium participation and overarching principles. The upload of summary statistics for this working group will be as listed in 7.1 and not to the Helmholtz servers; this analysis plan therefore intentionally supplements information in this document and deviates from it in this specific regard.

The coordination and leadership of this work is supported by UKRI (UK) Future Leaders Fellowship (MR/ S016538/1 and MR/S016538/2) to Watt, Imperial College London, UK. The work is supported by the GO consortium, led by Zeggini (Germany). The analysis lead is McDonald (Birmingham, USA). The analysis lead is based at the Central Analysis Site and will oversee all aspects of the analysis and meta-analysis, supported by the group. Any funder(s) of the study were not involved in the study design; collection, analysis, and interpretation of data; writing of the report; or decision to submit the paper for publication.

## Data Availability

No data has been produced

## Acknowledgments

We acknowledge the GO-PTOA working group’s members for their contribution to this work by representing cohorts and/or contributing methodological expertise. Contributing cohorts, institutions and working group members are listed on page 3 of the document (https://www.genetics-osteoarthritis.com/active-working-groups/gwas-meta-analysis-of-post-traumatic-osteoarthritis-of-the-knee/index.html). Additional acknowledgements specific to cohorts are listed below.

## VA Million Veteran Program (MVP) Acknowledgements

MVP Executive Committee

- Co-Chair: J. Michael Gaziano, M.D., M.P.H. VA Boston Healthcare System, 150 S. Huntington Avenue, Boston, MA 02130
- Co-Chair: Sumitra Muralidhar, Ph.D. US Department of Veterans Affairs, 810 Vermont Avenue NW, Washington, DC 20420
- Rachel Ramoni, D.M.D., Sc.D., Chief VA Research and Development Officer US Department of Veterans Affairs, 810 Vermont Avenue NW, Washington, DC 20420
- Jean Beckham, Ph.D. Durham VA Medical Center, 508 Fulton Street, Durham, NC 27705
- Kyong-Mi Chang, M.D. Philadelphia VA Medical Center, 3900 Woodland Avenue, Philadelphia, PA 19104
- Christopher J. O’Donnell, M.D., M.P.H. VA Boston Healthcare System, 150 S. Huntington Avenue, Boston, MA 02130
- Philip S. Tsao, Ph.D. VA Palo Alto Health Care System, 3801 Miranda Avenue, Palo Alto, CA 94304
- James Breeling, M.D., Ex-Officio US Department of Veterans Affairs, 810 Vermont Avenue NW, Washington, DC 20420
- Grant Huang, Ph.D., Ex-Officio US Department of Veterans Affairs, 810 Vermont Avenue NW, Washington, DC 20420
- Juan P. Casas, M.D., Ph.D., Ex-Officio VA Boston Healthcare System, 150 S. Huntington Avenue, Boston, MA 02130

MVP Program Office

- Sumitra Muralidhar, Ph.D. US Department of Veterans Affairs, 810 Vermont Avenue NW, Washington, DC 20420
- Jennifer Moser, Ph.D. US Department of Veterans Affairs, 810 Vermont Avenue NW, Washington, DC 20420

MVP Recruitment/Enrollment

- Recruitment/Enrollment Director/Deputy Director, Boston – Stacey B. Whitbourne, Ph.D.; Jessica V. Brewer, M.P.H. VA Boston Healthcare System, 150 S. Huntington Avenue, Boston, MA 02130

MVP Coordinating Centers

- Clinical Epidemiology Research Center (CERC), West Haven – Mihaela Aslan, Ph.D.
- West Haven VA Medical Center, 950 Campbell Avenue, West Haven, CT 06516
- Cooperative Studies Program Clinical Research Pharmacy Coordinating Center, Albuquerque – Todd Connor, Pharm.D.; Dean P. Argyres, B.S., M.S.
- New Mexico VA Health Care System, 1501 San Pedro Drive SE, Albuquerque, NM 87108

Genomics Coordinating Center, Palo Alto – Philip S. Tsao, Ph.D.

○ VA Palo Alto Health Care System, 3801 Miranda Avenue, Palo Alto, CA 94304
○ MVP Boston Coordinating Center, Boston-J. Michael Gaziano, M.D., M.P.H.
○ VA Boston Healthcare System, 150 S. Huntington Avenue, Boston, MA 02130
○ MVP Information Center, Canandaigua – Brady Stephens, M.S.
○ Canandaigua VA Medical Center, 400 Fort Hill Avenue, Canandaigua, NY 14424

- VA Central Biorepository, Boston – Mary T. Brophy M.D., M.P.H.; Donald E. Humphries, Ph.D.; Luis E. Selva, Ph.D.
- VA Boston Healthcare System, 150 S. Huntington Avenue, Boston, MA 02130
- MVP Informatics, Boston – Nhan Do, M.D.; Shahpoor (Alex) Shayan, M.S.
- VA Boston Healthcare System, 150 S. Huntington Avenue, Boston, MA 02130
- MVP Data Operations/Analytics, Boston – Kelly Cho, M.P.H., Ph.D.
- VA Boston Healthcare System, 150 S. Huntington Avenue, Boston, MA 02130
- Director of Regulatory Affairs – Lori Churby, B.S.
- VA Palo Alto Health Care System, 3801 Miranda Avenue, Palo Alto, CA 94304

MVP Science

- Science Operations – Christopher J. O’Donnell, M.D., M.P.H. VA Boston Healthcare System, 150 S. Huntington Avenue, Boston, MA 02130
- Genomics Core – Christopher J. O’Donnell, M.D., M.P.H.; Saiju Pyarajan Ph.D. VA Boston Healthcare System, 150 S. Huntington Avenue, Boston, MA 02130
- Philip S. Tsao, Ph.D. VA Palo Alto Health Care System, 3801 Miranda Avenue, Palo Alto, CA 94304
- Data Core – Kelly Cho, M.P.H, Ph.D. VA Boston Healthcare System, 150 S. Huntington Avenue, Boston, MA 02130
- VA Informatics and Computing Infrastructure (VINCI) – Scott L. DuVall, Ph.D. VA Salt Lake City Health Care System, 500 Foothill Drive, Salt Lake City, UT 84148
- Data and Computational Sciences – Saiju Pyarajan, Ph.D. VA Boston Healthcare System, 150 S. Huntington Avenue, Boston, MA 02130
- Statistical Genetics – Elizabeth Hauser, Ph.D. Durham VA Medical Center, 508 Fulton Street, Durham, NC 27705
- Yan Sun, Ph.D. Atlanta VA Medical Center, 1670 Clairmont Road, Decatur, GA 30033
- Hongyu Zhao, Ph.D. West Haven VA Medical Center, 950 Campbell Avenue, West Haven, CT 06516

Current MVP Local Site Investigators

- Atlanta VA Medical Center (Peter Wilson, M.D.) 1670 Clairmont Road, Decatur, GA 30033
- Bay Pines VA Healthcare System (Rachel McArdle, Ph.D.) 10,000 Bay Pines Blvd Bay Pines, FL 33744
- Birmingham VA Medical Center (Louis Dellitalia, M.D.) 700 S. 19th Street, Birmingham AL 35233
- Central Western Massachusetts Healthcare System (Kristin Mattocks, Ph.D., M.P.H.) 421 North Main Street, Leeds, MA 01053
- Cincinnati VA Medical Center (John Harley, M.D., Ph.D.) 3200 Vine Street, Cincinnati, OH 45220
- Clement J. Zablocki VA Medical Center (Jeffrey Whittle, M.D., M.P.H.) 5000 West National Avenue, Milwaukee, WI 53295
- VA Northeast Ohio Healthcare System (Frank Jacono, M.D.) 10701 East Boulevard, Cleveland, OH 44106
- Durham VA Medical Center (Jean Beckham, Ph.D.) 508 Fulton Street, Durham, NC 27705
- Edith Nourse Rogers Memorial Veterans Hospital (John Wells., Ph.D.) 200 Springs Road, Bedford, MA 01730
- Edward Hines, Jr. VA Medical Center (Salvador Gutierrez, M.D.) 5000 South 5th Avenue, Hines, IL 60141
- Veterans Health Care System of the Ozarks (Gretchen Gibson, D.D.S., M.P.H.) 1100 North College Avenue, Fayetteville, AR 72703
- Fargo VA Health Care System (Kimberly Hammer, Ph.D.) 2101 N. Elm, Fargo, ND 58102
- VA Health Care Upstate New York (Laurence Kaminsky, Ph.D.) 113 Holland Avenue, Albany, NY 12208
- New Mexico VA Health Care System (Gerardo Villareal, M.D.) 1501 San Pedro Drive, S.E. Albuquerque, NM 87108
- VA Boston Healthcare System (Scott Kinlay, M.B.B.S., Ph.D.) 150 S. Huntington Avenue, Boston, MA 02130
- VA Western New York Healthcare System (Junzhe Xu, M.D.) 3495 Bailey Avenue, Buffalo, NY 14215-1199
- Ralph H. Johnson VA Medical Center (Mark Hamner, M.D.) 109 Bee Street, Mental Health Research, Charleston, SC 29401
- Columbia VA Health Care System (Roy Mathew, M.D.) 6439 Garners Ferry Road, Columbia, SC 29209
- VA North Texas Health Care System (Sujata Bhushan, M.D.) 4500 S. Lancaster Road, Dallas, TX 75216
- Hampton VA Medical Center (Pran Iruvanti, D.O., Ph.D.) 100 Emancipation Drive, Hampton, VA 23667
- Richmond VA Medical Center (Michael Godschalk, M.D.) 1201 Broad Rock Blvd., Richmond, VA 23249
- Iowa City VA Health Care System (Zuhair Ballas, M.D.) 601 Highway 6 West, Iowa City, IA 52246-2208
- Eastern Oklahoma VA Health Care System (Douglas Ivins, M.D.) 1011 Honor Heights Drive, Muskogee, OK 74401
- James A. Haley Veterans’ Hospital (Stephen Mastorides, M.D.) 13000 Bruce B. Downs Blvd, Tampa, FL 33612
- James H. Quillen VA Medical Center (Jonathan Moorman, M.D., Ph.D.) Corner of Lamont & Veterans Way, Mountain Home, TN 37684
- John D. Dingell VA Medical Center (Saib Gappy, M.D.) 4646 John R Street, Detroit, MI 48201
- Louisville VA Medical Center (Jon Klein, M.D., Ph.D.) 800 Zorn Avenue, Louisville, KY 40206
- Manchester VA Medical Center (Nora Ratcliffe, M.D.) 718 Smyth Road, Manchester, NH 03104
- Miami VA Health Care System (Hermes Florez, M.D., Ph.D.) 1201 NW 16th Street, 11 GRC, Miami FL 33125
- Michael E. DeBakey VA Medical Center (Olaoluwa Okusaga, M.D.) 2002 Holcombe Blvd, Houston, TX 77030
- Minneapolis VA Health Care System (Maureen Murdoch, M.D., M.P.H.) One Veterans Drive, Minneapolis, MN 55417
- N. FL/S. GA Veterans Health System (Peruvemba Sriram, M.D.) 1601 SW Archer Road, Gainesville, FL 32608
- Northport VA Medical Center (Shing Shing Yeh, Ph.D., M.D.) 79 Middleville Road, Northport, NY 11768
- Overton Brooks VA Medical Center (Neeraj Tandon, M.D.) 510 East Stoner Ave, Shreveport, LA 71101
- Philadelphia VA Medical Center (Darshana Jhala, M.D.) 3900 Woodland Avenue, Philadelphia, PA 19104
- Phoenix VA Health Care System (Samuel Aguayo, M.D.) 650 E. Indian School Road, Phoenix, AZ 85012
- Portland VA Medical Center (David Cohen, M.D.) 3710 SW U.S. Veterans Hospital Road, Portland, OR 97239
- Providence VA Medical Center (Satish Sharma, M.D.) 830 Chalkstone Avenue, Providence, RI 02908
- Richard Roudebush VA Medical Center (Suthat Liangpunsakul, M.D., M.P.H.) 1481 West 10th Street, Indianapolis, IN 46202
- Salem VA Medical Center (Kris Ann Oursler, M.D.) 1970 Roanoke Blvd, Salem, VA 24153
- San Francisco VA Health Care System (Mary Whooley, M.D.) 4150 Clement Street, San Francisco, CA 94121
- South Texas Veterans Health Care System (Sunil Ahuja, M.D.) 7400 Merton Minter Boulevard, San Antonio, TX 78229
- Southeast Louisiana Veterans Health Care System (Joseph Constans, Ph.D.) 2400 Canal Street, New Orleans, LA 70119
- Southern Arizona VA Health Care System (Paul Meyer, M.D., Ph.D.) 3601 S 6th Avenue, Tucson, AZ 85723
- Sioux Falls VA Health Care System (Jennifer Greco, M.D.) 2501 W 22nd Street, Sioux Falls, SD 57105
- St. Louis VA Health Care System (Michael Rauchman, M.D.) 915 North Grand Blvd, St. Louis, MO 63106
- Syracuse VA Medical Center (Richard Servatius, Ph.D.) 800 Irving Avenue, Syracuse, NY 13210
- VA Eastern Kansas Health Care System (Melinda Gaddy, Ph.D.) 4101 S 4th Street Trafficway, Leavenworth, KS 66048
- VA Greater Los Angeles Health Care System (Agnes Wallbom, M.D., M.S.) 11301 Wilshire Blvd, Los Angeles, CA 90073
- VA Long Beach Healthcare System (Timothy Morgan, M.D.) 5901 East 7th Street Long Beach, CA 90822
- VA Maine Healthcare System (Todd Stapley, D.O.) 1 VA Center, Augusta, ME 0433
- VA New York Harbor Healthcare System (Scott Sherman, M.D., M.P.H.) 423 East 23rd Street, New York, NY 10010
- VA Pacific Islands Health Care System (George Ross, M.D.) 459 Patterson Rd, Honolulu, HI 96819
- VA Palo Alto Health Care System (Philip Tsao, Ph.D.) 3801 Miranda Avenue, Palo Alto, CA 94304-1290
- VA Pittsburgh Health Care System (Patrick Strollo, Jr., M.D.) University Drive, Pittsburgh, PA 15240
- VA Puget Sound Health Care System (Edward Boyko, M.D.) 1660 S. Columbian Way, Seattle, WA 98108-1597
- VA Salt Lake City Health Care System (Laurence Meyer, M.D., Ph.D.) 500 Foothill Drive, Salt Lake City, UT 84148
- VA San Diego Healthcare System (Samir Gupta, M.D., M.S.C.S.) 3350 La Jolla Village Drive, San Diego, CA 92161
- VA Sierra Nevada Health Care System (Mostaqul Huq, Pharm.D., Ph.D.) 975 Kirman Avenue, Reno, NV 89502
- VA Southern Nevada Healthcare System (Joseph Fayad, M.D.) 6900 North Pecos Road, North Las Vegas, NV 89086
- VA Tennessee Valley Healthcare System (Adriana Hung, M.D., M.P.H.) 1310 24th Avenue, South Nashville, TN 37212
- Washington DC VA Medical Center (Jack Lichy, M.D., Ph.D.) 50 Irving St, Washington, D. C. 20422
- W.G. (Bill) Hefner VA Medical Center (Robin Hurley, M.D.) 1601 Brenner Ave, Salisbury, NC 28144
- White River Junction VA Medical Center (Brooks Robey, M.D.) 163 Veterans Drive, White River Junction, VT 05009
- William S. Middleton Memorial Veterans Hospital (Robert Striker, M.D., Ph.D.) 2500 Overlook Terrace, Madison, WI 53705

## UK Biobank (UKB) Acknowledgements

This research will include data from the UK Biobank (UKB) resource. We thank UKB and its participants. This work will use data provided by patients and collected by NHS as part of their care and support. UKB has approval from the North West Multi-centre Research Ethics Committee (MREC), Haydock, UK as a Research Tissue Bank (RTB). Use of UKB data for this project received UKB approvals in June 2020 (project number 52507). The publication does not represent the views of the NHS, NIHR or the Department of Health.

## Estonian Biobank (EstBB) Acknowledgements

Andres Metspalu, Lili Milani, Tõnu Esko, Reedik Mägi, Mari Nelis, Georgi Hudjashov

## FUNDING

### Imperial College London funding and coordination of the group

FW and RK are funded by a UK Research and Innovation (UKRI) Future Leaders Fellowship to FW, (MR/S016538/1 and MR/S016538/2).

### Oxford funding

LJD is supported by Wellcome Trust grant 208750/Z/17/Z.

### VA Million Veteran Program (MVP) funding

This research is based on data from the Million Veteran Program, Office of Research and Development, Veterans Health Administration, and was supported by award #I01RX002745. This publication does not represent the views of the Department of Veteran Affairs or the United States Government.

### Estonian Biobank (EstBB) funding

This study was funded by European Union through the European Regional Development Fund Project No. 2014-2020.4.01.15-0012 GENTRANSMED and the Estonian Research Council grant PUT (PRG1911). Data analysis was carried out in part in the High-Performance Computing Center of University of Tartu. The activities of the EstBB are regulated by the Human Genes Research Act, which was adopted in 2000 specifically for the operations of the EstBB. Individual level data analysis in the EstBB was carried out under ethical approval [1.1-12/624] from the Estonian Committee on Bioethics and Human Research (Estonian Ministry of Social Affairs), using data according to release application [N05] from the Estonian Biobank

### University of Iceland

The research was supported by University of Iceland. No other authors declare a funding.

### APPENDIX

#### Appendix 1 Inclusion criteria for knee PTOA^1^

##### Appendix 1A Suggested approach to knee injury case definition for time-dependent analyses, including time-to-event PTOA cases

Appendix 1A: Suggested approach to knee injury case definition for time-dependent analyses, including time-to-event PTOA cases
| Structure | Location | Trauma | Supporting Description |
| --- | --- | --- | --- |
| 1 | 1 | 1 | Evidence of an acute knee injury or a surgical procedure which is highly suggestive of this (this includes any clinically significant/ injury to the structures of the joint tissues or periarticular bone*, and/or dislocation of the joint) |
| 1 | 1 | 0 | Evidence of a clinically significant knee joint abnormality which is consistent with injury or a procedure on the knee which implies this, without definite evidence of trauma |
| 1 | 0 | 1 | Evidence of a clinically significant acute joint injury or procedure which is suggestive of this (but not specific to knee) |
| 1 | 0 | 0 | Evidence of a clinically significant joint abnormality or procedure on a joint which implies this (but not specific to knee), without definite evidence of trauma |
| 0 | 1 | 1 | Evidence of a traumatic injury involving the knee, but without definite evidence of clinical significance |
| 0 | 1 | 0 | Evidence of knee involvement without definite evidence of trauma or clinical significance |
| 0 | 0 | 1 | Evidence of traumatic episode compatible with a possible knee joint injury without definite evidence of knee involvement or clinical significance to the knee |
| 0 | 0 | 0 | No evidence of any of the above |

###### Structure, location and trauma codes supporting case ascertainment of acute knee injury

*To be included, cases should achieve a code for structure, location and trauma in a set timeframe, either from a single code or in combination, see **Inclusion code lists**^1^ referencing these categories, including lists of ‘1,1,1’ codes (single codes which are sufficient for inclusion if preferred).

Internal structures of the knee are intra-articular structures of the joint tissues include peri-articular bone, and/or dislocation of the joint, or surgical procedures carried out on these structures in keeping with injury to them e.g. reconstruction of a ligament).

Examples lacking sufficient evidence of clinically significant injury to an internal structure include MCL sprains (as opposed to rupture); presence of synovial plica; patello-femoral subluxation (as opposed to dislocation); quadriceps and patellar tendon injuries. Examples lacking sufficient evidence for acute trauma e.g. contusion, superficial wound, knee sprain.

##### Appendix 1B Known ‘Case-code PTOA’ codes for consideration in non-time dependent analyses

To be included, cases should achieve a code sufficient for osteoarthritis, location (i.e. knee) and linked prior trauma (post-traumatic OA code), from a single code^1^ or in combination (e.g. knee location code can be separate). The identified examples are given below:

| ICD-10 Code | Description |
| --- | --- |
| M17.2 | post-traumatic gonarthrosis, bilateral |
| M17.3 | other post-traumatic gonarthrosis |
| M12.569* | traumatic arthropathy, unspecified knee |
| M19.16* | post-traumatic OA lower leg/knee |

\*These codes may only be applicable in certain datasets with these subcodes, e.g. US datasets. M12.5 and M19.1 are not sufficient for inclusion.
\*Cross mapping to ICD-9= 716.16, Traumatic arthropathy, lower leg

#### Appendix 2 Exclusion criteria for knee PTOA cases

##### Appendix 2A Suggested Exclusion criteria for time-dependent analyses^2^

###### Exclusion criteria

The presence of any of the following, diagnosed at **any time PRIOR** to the date of the knee injury^#^:

1. Osteoarthritis (or non-specified arthropathy which could be osteoarthritis^a^) of either knee, hip or ankle
2. Partial or total knee arthroplasty of either knee, hip or ankle
3. Clinically significant injuries to the hip including hip and pelvic fracture.

The presence of any of the following, diagnosed **6 months or more PRIOR** to the date of the knee injury^#^:

4. Chronic instability or recurrent patellofemoral dislocation of the knee without evidence of a prior acute knee injury of known timing
5. Acquired structural abnormalities of the knee, without evidence of a prior acute knee injury of known timing (e.g. degenerative meniscus included cystic change, chondrocalcinosis) or which are not attributable to an acute knee injury that meets the inclusion criteria^b^.

The presence of any of the following **at ANY TIME** (prior, during or after the date of knee injury^#^ and up until the first date of osteoarthritis or equivalent):

6. Inflammatory arthritis^C^ or associated connective tissue disorder (e.g. rheumatoid arthritis, psoriatic arthritis, juvenile idiopathic arthritis, enteropathic arthritis, inflammatory spondyloarthropathy, Lupus or Sjogren’s syndrome; generalised or lower limb gout, CPPD, reactive arthritis or septic arthritis); neuropathic arthritis or inherited collagen vascular disease affecting joints.
7. Clinically significant congenital structural abnormality of the knee, or clinically significant congenital or acquired structural abnormality of the hip or ankle (e.g. discoid meniscus, absence of anterior cruciate ligament, congenital dislocation of hip, protrusio acetabuli)
8. Septic arthritis or osteomyelitis of the knee
9. Individuals with non-normal weight bearing (e.g. wheelchair user, those with spina bifida or cerebral palsy, lower limb amputations)

Evidence of more extensive musculoskeletal trauma **within the ‘injury window’** for the acute knee injury^#^:

10. Open or closed fractures of the lower limb which are not likely to be intra-articular in origin (e.g. proximal or mid femoral fractures, proximal or distal tibial fractures, distal fibular fractures).
11. Clinically significant neurovascular or nerve palsies of the lower limb likely caused by lower limb trauma e.g. peroneal nerve, sciatic nerve or femoral nerve palsies.
12. Multisite musculoskeletal trauma, outside of the knee.

^#^where the date of the knee injury is the start of a three month ‘injury window’, which begins at the first evidence of injury

^a^ This includes climacteric arthritis, HPOA, chondrolysis and inherited or metabolic disorders which always give rise to OA

^b^ Osgood Schlatters disease (osteochondritis of the tibial tuberosity) and functional disorders of the patella including chondromalacia patellae are not sufficient to fulfil this criterion. Operative procedures to the joint such as arthroscopy are not sufficient to meet this criterion, unless definitely attributable to a relevant structural abnormality which was not due to an acute injury.

^c^Allergic arthritis, transient arthritis, palindromic arthritis, post immunisation arthritis, intermittent hydrarthrosis are not considered sufficient/specific.

See **Exclusion code lists**^2^ for ICD-10 codes

##### Appendix 2B Suggested modified Exclusion criteria (for non-time-dependent analyses)^3^

Evidence of any of the following:

1. Inflammatory arthritis^C^ or associated connective tissue disorder (e.g. rheumatoid arthritis, psoriatic arthritis, juvenile idiopathic arthritis, enteropathic arthritis, inflammatory spondyloarthropathy, Lupus or Sjogren’s syndrome; generalised or lower limb gout, CPPD, reactive arthritis or septic arthritis); neuropathic arthritis or inherited collagen vascular disease affecting joints.
2. Clinically significant congenital structural abnormality of the knee (e.g. discoid meniscus, absence of anterior cruciate ligament)
3. Septic arthritis or osteomyelitis of the knee
4. Non-normal weight bearing (e.g. wheelchair user, those with spina bifida or cerebral palsy, lower limb amputations)
5. Open or closed fractures of the lower limb which are not likely to be intra-articular in origin (e.g. proximal or mid femoral fractures, proximal or distal tibial fractures, distal fibular fractures).
6. Clinically significant neurovascular or nerve palsies of the lower limb likely caused by lower limb trauma e.g. peroneal nerve, sciatic nerve or femoral nerve palsies.
7. Multisite musculoskeletal trauma, outside of the knee.

Please note these are <u>exclusions 6-12</u> of the full exclusion criteria listed in Appendix 2A.

See **Exclusion code lists** for this subset of the ICD-10 codes (categories 6-12) and for mapped ICD-9 codes^3^

In addition, for (i) ‘time to event’ PTOA cases (where the presence of both injury codes and OA codes are used to define PTOA cases for case control analyses): it is important that those with knee OA occurring <u>before</u> the injury, i.e. exclusion criterion 1 from Appendix 2A, are excluded. That is:

1. Osteoarthritis (or non-specified arthropathy which could be osteoarthritis) of either knee, hip or ankle

#### Appendix 3 Definitions of knee OA

##### Appendix 3A Knee OA outcomes after injury^4^

Outcomes selected include categories 1 and 2 using the OA classification in Table below. In some cases, category 2 codes do require a knee specific code (which in all cases makes them a 1) to sufficiently increase their specificity (primarily arthroplasty codes).

For time-dependent analyses, earliest available OA codes should always be used, i.e. revision arthroplasty codes would be superseded by an earlier first arthroplasty code; arthroplasty codes would be superseded by an earlier non arthroplasty OA code. Revision codes suggesting there has been prior surgery have been specifically highlighted as an earlier code would be expected, although they can act as OA outcome codes where no others exist*.

^^^Column ‘Knee/hip code’ – where blank is no action and ‘yes’ indicates that code requires a knee code to ensure sufficient specificity (relating to arthroplasty codes).

*Column ‘Look for earlier code as knee revision surgery’ - where blank is no action and 1 notes revision code, i.e. where there should be an earlier OA outcome code as this is repeated surgery

See associated adjudication of **OA outcome code** lists^4^ eg ICD10.

Please note that PTOA knee codes <u>should</u> be included in these lists for time dependent analyses.

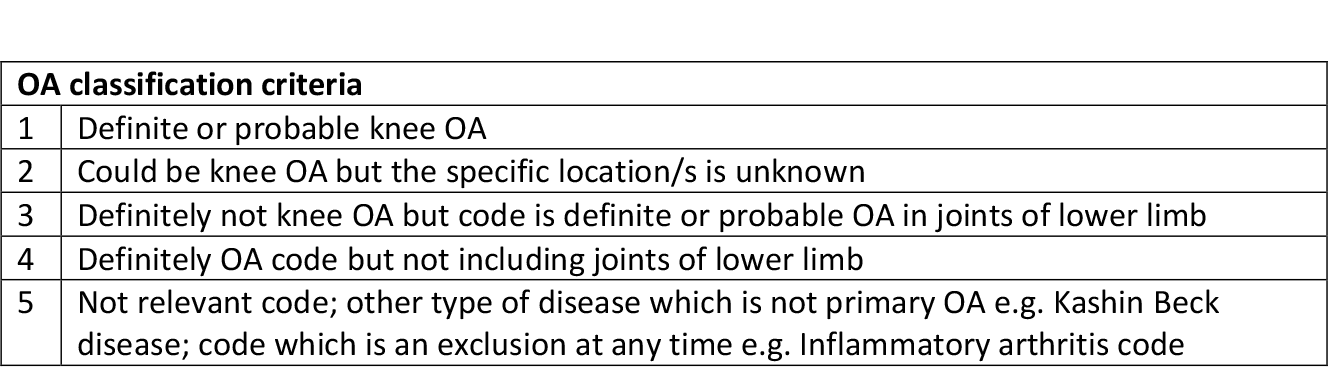

##### Appendix 3B Definition of idiopathic knee OA comparator group^5^

Idiopathic knee OA codes (KIOA) need <u>higher specificity</u> than those listed in 3A as OA outcomes.

These codes are a <u>subset</u> of ALLKOA, i.e. all the ‘category 1’ knee OA outcome code lists e.g. ICD10.

KIOA include cases of definite or probable knee OA, without prior knee trauma or ever history of PTOA

- All category 1 codes from the outcome code list given below (ALLKOA)
- Removing all secondary and ‘case code PTOA’ (CCPTOA), as listed in Appendix 1B.
- Removing all cases with history of prior knee injury as defined in Appendix 1A.

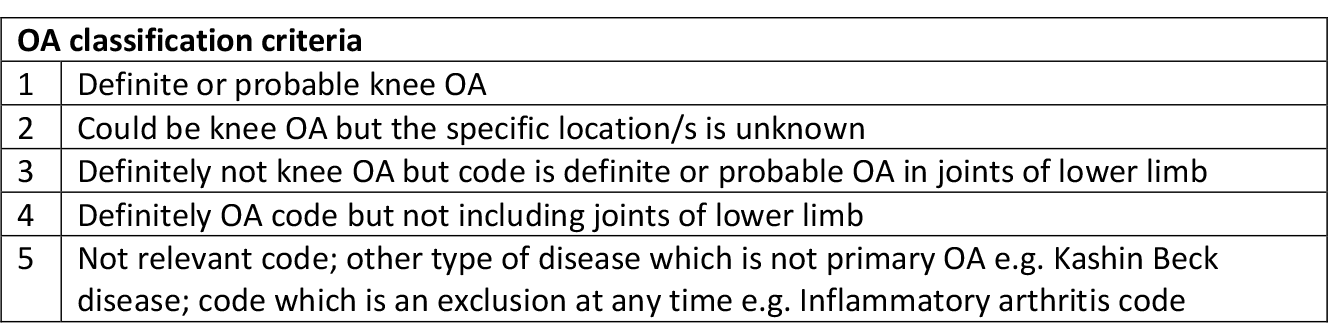

Suggested ‘idiopathic knee OA’ ICD-10 code list:

M170

M171

M179

Suggested ‘idiopathic knee OA’ ICD-9 code list:

715.16

715.36

715.96

#### Appendix 4 Summary of inclusion and exclusion criteria for each GWAS analysis

Please note this has now been superseded by new Appendix 5.

#### Appendix 5 GO-PTOA Primary Analysis including detailed methods for prioritised case-control analyses

##### 2.0. Research questions

Our main question is to determine whether there is genetic variation associated with the development of knee PTOA.

Our overall hypothesis is that there is genetic variation associated with the development of knee PTOA. We want to definitively answer whether these variants are the same or different from idiopathic knee OA (iOA) genetic variants. Under this overarching hypothesis, we will execute the following aims:

###### Aim 1 Identify genetic variation associated with knee PTOA independent of knee iOA

To achieve this aim, each cohort will perform a case-control GWAS of knee PTOA vs. knee iOA cases (without knee injury history).

###### Aim 2 Identify genetic variation significantly predictive of the onset of knee PTOA

Each cohort with available data will perform a time-to-event GWAS of all knee injured cases with available time information, looking at the subsequent event of presence of knee OA/PTOA, to 20 years of follow up or censor.

###### Aim 3 Identify genetic variation associated with the onset of knee PTOA compared with knee injured controls without OA

Each cohort with available data will perform a case-control GWAS of knee PTOA cases vs. knee injured controls (no OA with adequate follow up).

###### Aim 4 Identify genetic variation associated with knee PTOA compared with controls over 40 years of age without injury or OA

Each cohort with available data will perform:

A. case-control GWAS of knee PTOA cases versus Injury free, OA free controls;
B. case-control GWAS of knee injured versus injury free, OA free controls; and
C. case-control GWAS of knee iOA vs. Injury free, OA free controls

Results from these three models will be compared to identify genetic variants likely to be associated with knee PTOA rather than knee injury risk or knee iOA.

**Figure 1:**
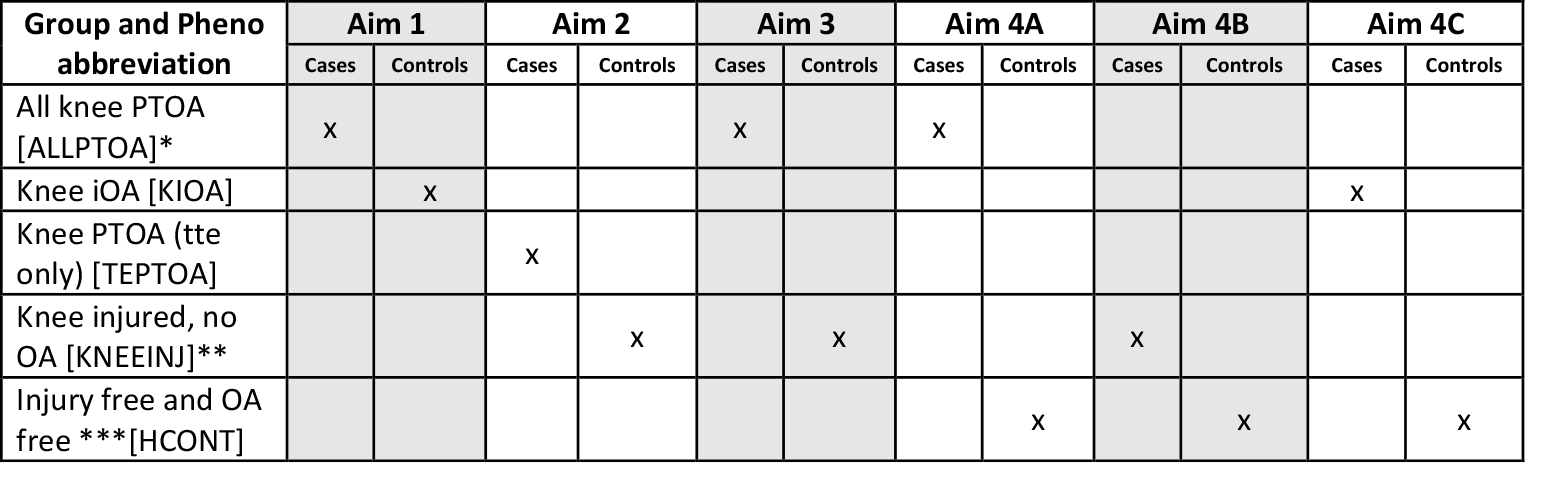
Summary of coding groups demonstrating redundance of use in Aims to facilitate preparation of phenotype files. Additional details regarding each group are provided in section 3.0. *ALLPTOA = time to event PTOA (TEPTOA) + case code PTOA (CCPTOA) **based on minimum follow up. NB For those who have defined ‘time to event Knee Injured controls’ in aim 2, please note the Knee injured cases in aim 4B are slightly different (as OA at any time is excluded, not just up to 20 years). ***based on minimum age

Below is a flow diagram to illustrate the order in which cohorts should work in. Please refer to this diagram to ensure that you have the relevant sign off from coordinating & analysis site at the appropriate time prior to running individual GWAS.

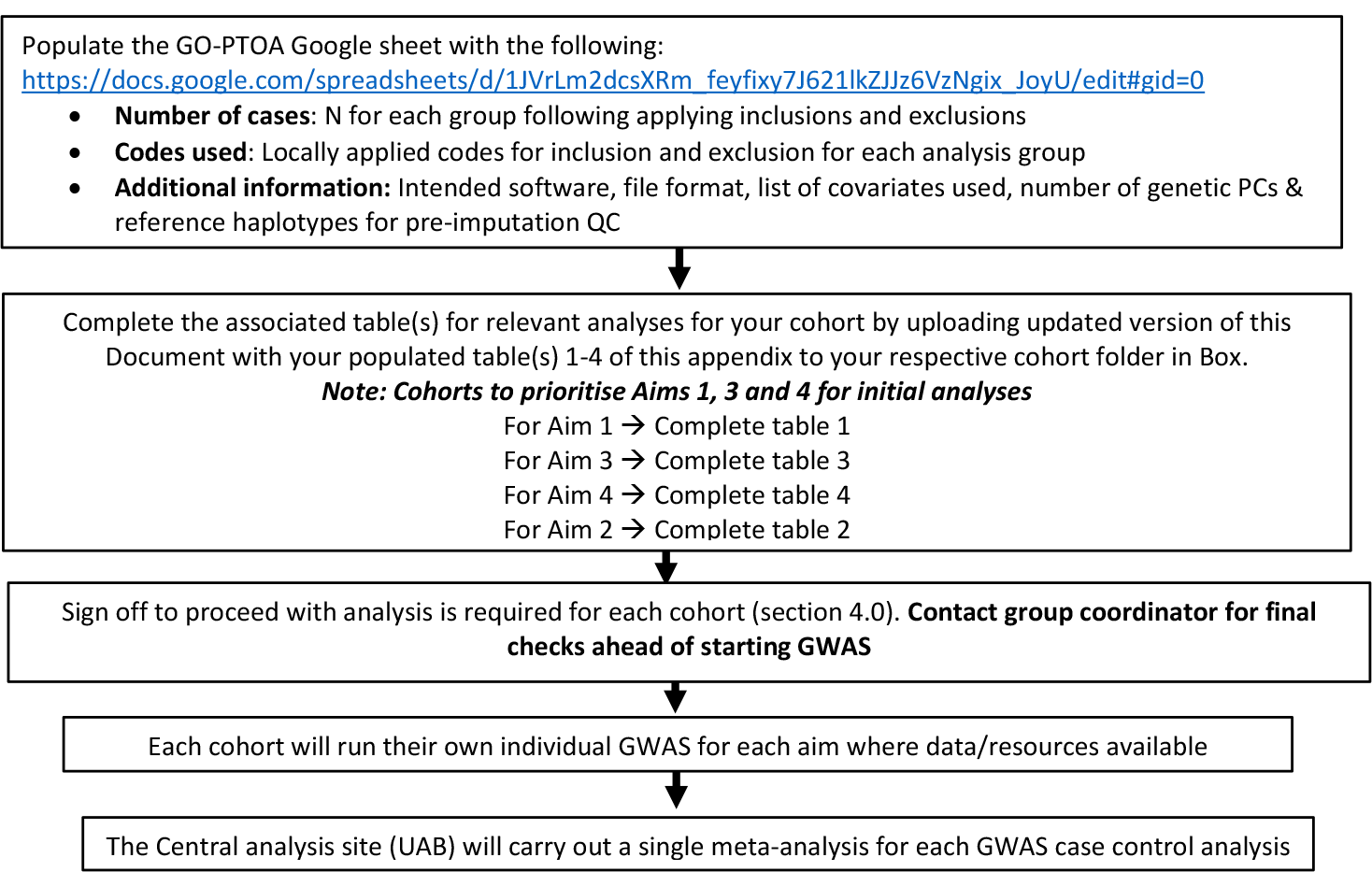

##### 3.0. Phenotype Definition by Analysis Aim

**The group has prioritised Aims 1, 3 and 4 for initial analyses. Aim 2 methods will be included in a separate appendix**.

**Aim 1: Identify genetic variation associated with knee PTOA independent of knee iOA**

###### Inclusions

- Evidence of at least one clinically significant knee injury in a participant which is likely due to acute trauma to an internal joint structure (at/within the knee joint) rather than elsewhere (Appendix 1A) AND a subsequent OA code which is likely of the same knee (or of either knee if side is not known) at least 6 months after injury (Appendix 3A). (When considering evidence for a clinically significant knee injury, this can be a range of knee injuries, but ideally without evidence of more extensive trauma, see Eligibility criteria, Appendix 1 & 2)
- Injury occurring between the ages of **16** (or 18 if lack of consent for this) **and 50 inclusive**

###### AND/OR

- Evidence of a specific diagnosis by PTOA code (see Appendix 1B), <u>where not identified already</u> using the inclusion approach above (Appendix 1A).

###### Exclusions

- Where there is time information, consider applying exclusions as per Appendix 2A if feasible

NB Pre-existing OA prior to the injury, or conditions which suggest this, <u>should always be excluded</u>

- OA code within 6 months or less after first injury code, where this is available
- For all cases (including where identified by specific PTOA code diagnosis), simplified exclusions, defined by cohort, supported by Appendix 2B, should be applied

###### Inclusions

- Probable idiopathic knee OA <u>at any time</u> (see Appendix 3B, i.e. ‘category 1’ knee OA from 3A, removing all post-traumatic knee OA codes) (NB when reporting this group’s characteristics, the first eligible OA code for this should be used)

###### Exclusions

- <u>Evidence of an acute knee injury</u> **<u>at any time</u>**<u>(i.e. exclude</u> all definite acute knee injury, defined by cohort as supported by Appendix 1A and related inclusion codes)
- A PTOA knee code at any time (Appendix 1B)
- Other simplified exclusions, defined by cohort, as supported by Appendix 2B

Please complete the below table to summarize the cases and controls included in Aim 1 analyses from your study. In addition, please populate the GO-PTOA Google sheet (https://docs.google.com/spreadsheets/d/1JVrLm2dcsXRm_feyfixy7J621lkZJJz6VzNgix_JoyU/edit#gid=1072245856) with: 1) N for each group; 2) locally applied codes for inclusion for each group and; 3) exclusion codes for each group.

Contact the group coordinator for final checks ahead of starting each GWAS.

Upload your GWAS for Aim 1 to folder **‘Aim1_ALLPTOA_vs_KIOA’** on Box. Please include ‘Aim1_ALLPTOA_vs_KIOA’ in the name of your file uploaded.

**Table 1.**
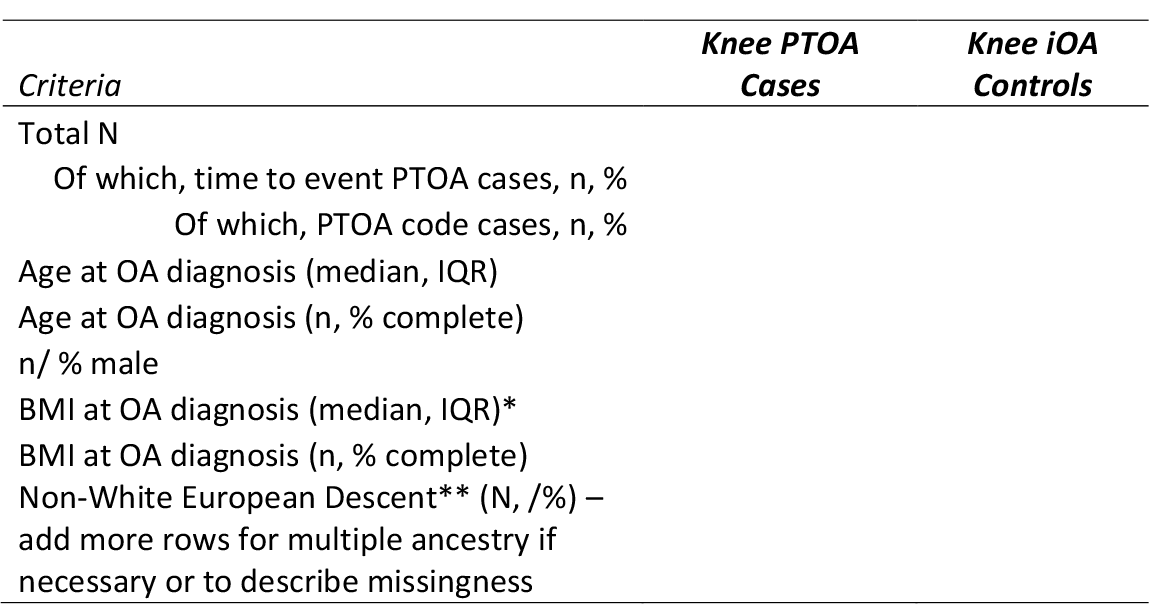
Aim 1 Descriptives. * BMI at time of PTOA or OA diagnosis (within 2 years of this) will used to describe the groups and also in the BMI sensitivity analyses (**see Section 5**). If not available at diagnosis, please specify the age/ time at which BMI is being reported. ** If significant number non-White European descent subjects please specify breakdown by major ancestry.

| *Criteria* | *Knee PTOA Cases* | *Knee iOA Controls* |
| --- | --- | --- |
| Total N |  |  |
| Of which, time to event PTOA cases, n, % |  |  |
| Of which, PTOA code cases, n, % |  |  |
| Age at OA diagnosis (median, IQR) |  |  |
| Age at OA diagnosis (n, % complete) |  |  |
| n/ % male |  |  |
| BMI at OA diagnosis (median, IQR)* |  |  |
| BMI at OA diagnosis (n, % complete) |  |  |
| Non-White European Descent** (N, /%) –<br>add more rows for multiple ancestry if<br>necessary or to describe missingness |  |  |

**Aim 2: Identify genetic variation significantly predictive of the onset of knee PTOA**

**<u>NOTE: Aim 2 is tabled pending additional discussion/methods. Case control analyses have been prioritized by group to be actioned first. Please move to Aim 3</u>**.

A time-to-event GWAS among the PTOA at risk population of participants with a history of knee injury. Knowledge that the injury predates the development of PTOA is necessary to perform this time to event analysis.

**Fig 2.**
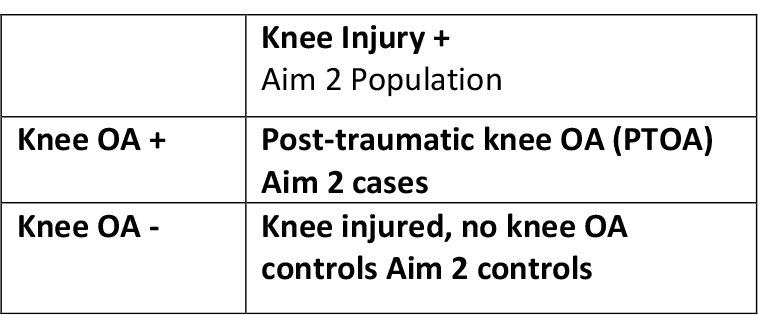
Comparison of general criteria considered to define PTOA of the knee within participants with history of a knee injury in Aim 2.

###### Inclusions for both Aim 2 cases and controls

Both cases and controls must have had a knee injury exposure defined by:

- A timed definite acute knee injury (defined by cohort as supported by Appendix 1A and related inclusion codes)
- Injury occurring between the ages of **16** (or 18 if lack of consent for this) **and 50 inclusive**

###### Exclusions

- Exclusions defined by cohort, as supported by Appendix 2A and related exclusion codes
- OA code within 6 months or less after first injury code
- Pre-existing OA prior to the injury, or conditions which suggest this, should always be excluded (as supported by Appendix 2A)

###### Outcomes

- Knee OA outcomes (defined by cohort, as supported by Appendix 3A, which can include PTOA knee codes (see appendix 1B)
- An observation period of 6 months to <u>20 years after the knee injury</u> should be applied for cases of subsequent OA, or censoring to that date (or at death or last record if sooner)
- Knee OA diagnosis in the 6 months after injury ascertainment should be excluded (as likely pre-existing OA)

Please populate the GO-PTOA Google sheet https://docs.google.com/spreadsheets/d/1JVrLm2dcsXRm_feyfixy7J621lkZJJz6VzNgix_JoyU/edit#gid=1072245856 with: 1) N for each group; 2) locally applied codes for inclusion and; 3) exclusion codes for each group. *Please complete table below if possible, noting if this analysis is not being carried out yet*.

Contact coordinator for final checks ahead of starting GWAS.

Upload your GWAS for Aim 2 to folder **‘Aim2_TEPTOA_vs_KNEEINJ’** on Box. Please include ‘Aim2_TEPTOA_vs_KNEEINJ’ in the name of your file uploaded.

**Table 2.** Aim 2 Descriptives. * If available for all cases and controls, record Age and BMI at time injury (or +/- 2 years of this). If only available for a subset, please report n, % completeness, as well as Age and BMI at time of injury or else at OA diagnosis (+/- 2 years). BMI will be used for description and for sensitivity analyses (**see Section 5**). ** If significant number non-White European descent subjects, please specify breakdown by major ancestry. Please state NA if not available.

| *Criteria* | *Time to Event*<br>*Knee PTOA*<br>*Cases* | *Knee injured, no OA Controls* |
| --- | --- | --- |
| N |  |  |
| Age at time of injury* (median, IQR) |  |  |
| Age at time of injury, complete (n, %) |  |  |
| Age at PTOA diagnosis (median, IQR) |  |  |
| Age at PTOA diagnosis, complete (n, %) |  |  |
| n/ % male |  |  |
| BMI at time of injury* (median, IQR) |  |  |
| BMI at time of injury (n, % complete) |  |  |
| BMI at PTOA diagnosis, or inclusion (median, IQR) |  |  |
| BMI at PTOA diagnosis, or inclusion (n, % complete) |  |  |
| Non-White European Descent** (n, %) – add more rows for multiple ancestry if necessary or to describe missingness |  |  |
| Follow up time after injury, years (median, IQR) |  |  |

**Aim 3: Identify genetic variation associated with the onset of knee PTOA**

###### Inclusions

- Evidence of at least one clinically significant knee injury in a participant*, which is likely due to acute trauma to an internal joint structure (at/within the knee joint) rather than elsewhere (Appendix 1A) AND a subsequent OA code which is likely of the same knee (or of either knee if side is not known) at least 6 months after injury code (Appendix 3A). (When considering evidence for a clinically significant knee injury, this can be a range of knee injuries, but ideally without evidence of more extensive trauma, see Eligibility criteria, Appendix 1 & 2)
- Knee injury occurring between the ages of **16** (or 18 if lack of consent for this) **and 50 inclusive**

###### AND/OR

- Evidence of a specific diagnosis by PTOA code (see Appendix 1B), <u>where not identified already</u> using the inclusion approach above (Appendix 1A).

###### Exclusions

- Where there is time information, consider applying exclusions as per Appendix 2A if feasible

NB Pre-existing OA prior to the injury, or conditions which suggest this, should <u>always be excluded</u>

- OA code within 6 months or less after injury code, where this is available
- For all cases (including where identified by specific PTOA code diagnosis), simplified exclusions, defined by cohort, supported by Appendix 2A or 2B, should be applied

###### Inclusions

- A definite acute knee injury (defined by cohort as supported by Appendix 1A and related inclusion codes)
- Injury occurring between the ages of **16** (or 18 if lack of consent for this) **and 50 inclusive**
- Evidence of at least 10 years follow up post knee injury is <u>recommended</u>

(If thresholding on 10 years of follow up is not possible, state local threshold, or else flag if not possible: still proceed, but this is relevant for sensitivity analysis^¥^)

###### Exclusions

- A knee OA or PTOA outcome code <u>at any time</u> (see appendix 3A and 1B)
- Exclusions defined by cohort, as supported by Appendix 2A/2B and related exclusion codes

Please populate the GO-PTOA Google sheet https://docs.google.com/spreadsheets/d/1JVrLm2dcsXRm_feyfixy7J621lkZJJz6VzNgix_JoyU/edit#gid=1072245856 with: 1) N for each group; 2) locally applied codes for inclusion and; 3) exclusion codes for each group. *Please complete table below if possible, noting if this analysis is not being carried out yet*.

Contact coordinator for final checks ahead of starting GWAS.

Upload your GWAS for Aim 3 to folder **‘Aim3_ALLPTOA_vs_KNEEINJ’** on Box. Please include ‘Aim3_ALLPTOA_vs_KNEEINJ’ in the name of your file uploaded.

**Table 3.** Aim 3 Descriptives. *If available in all cases and controls, please report and use Age and BMI at time of injury. BMI at time of injury will be used as a descriptor and in sensitivity analyses. Otherwise, please use BMI at time of PTOA or time of last follow up for table and BMI sensitivity analyses (**see Section 5**). ** If significant number non-White European descent subjects, please specify breakdown by major ancestry. Please state NA if not available.

|  | *Knee PTOA*<br>*Cases* | *Knee injured, no*<br>*OA Controls* |
| --- | --- | --- |
| *Criteria* |  |  |
| N |  |  |
| Of which, time to event PTOA cases, n, % |  |  |
| Of which, PTOA code cases, n, % |  |  |
| State local minimum follow up threshold for<br>Knee injured controls<sup>‡</sup>(10 years/7 years/5<br>years/median FU/no threshold) |  |  |
| Age at time of injury (median, IQR) |  |  |
| Age at time of injury, complete (n, %) |  |  |
| Age at time of PTOA diagnosis (median, IQR) |  |  |
| Age at time of PTOA diagnosis, complete (n,<br>%) |  |  |
| n/ % male |  |  |
| BMI at time of injury* (median, IQR) |  |  |
| BMI at time of injury, complete (n, %) |  |  |
| BMI at inclusion (PTOA diagnosis or time of<br>last follow up) (median, IQR) |  |  |
| BMI at inclusion, complete (n, %) |  |  |
| Non-White European Descent** (N)– add<br>more rows for multiple ancestry if necessary<br>or missingness |  |  |
| Follow up time after injury, years (median,<br>IQR) |  |  |

**Aim 4: Identify genetic variation associated with PTOA compared with age matched controls without injury or OA**

###### Inclusions

- Evidence of at least one clinically significant knee injury in a participant*, which is likely due to acute trauma to an internal joint structure (at/within the knee joint) rather than elsewhere (Appendix 1A) AND a subsequent OA code which is likely of the same knee (or of either knee if side is not known) at least 6 months after injury code (Appendix 3A). (When considering evidence for a clinically significant knee injury, this can be a range of knee injuries, but ideally without evidence of more extensive trauma, see Eligibility criteria, Appendix 1 & 2)
- Injury occurring between the ages of **16** (or 18 if lack of consent for this) **and 50 inclusive**

###### AND/OR

- Evidence of a specific diagnosis by PTOA code (see Appendix 1B), where not <u>identified already</u> using the inclusion approach above (Appendix 1A).

###### Exclusions

- Where there is time information, consider applying exclusions as per Appendix 2A if feasible

NB Pre-existing OA prior to the injury, or conditions which suggest this, should <u>always be excluded</u>

- OA code within 6 months or less after injury code, where this is available
- For all cases (including where identified by specific PTOA code diagnosis), simplified exclusions, defined by cohort, supported by Appendix 2A or 2B, should be applied

###### Inclusion

- Age 40 or more

###### Exclusions

- Evidence of an acute knee injury<u>at any time (</u>i.e. exclude all definite acute knee injury, defined by cohort as supported by Appendix 1A and related inclusion codes)
- An OA or PTOA outcome code <u>at any time (</u>see appendix 3A and 1B)
- Exclusions defined by cohort, as supported by Appendix 2A/2B and related exclusion codes

###### Inclusions

- A definite acute knee injury (defined by cohort as supported by Appendix 1A and related inclusion codes)
- Injury occurring between the ages of **16** (or 18 if lack of consent for this) **and 50 inclusive**
- Evidence of at least 10 years follow up post knee injury is recommended (If thresholding on 10 years of follow up is not possible, state local threshold in table below, or else flag if not possible: still proceed, but this is relevant for sensitivity analysis^¥^)

###### Exclusions

- An OA or PTOA outcome code <u>at any time (see Appendix 3A and 1B)</u>
- Exclusions defined by cohort, as supported by Appendix 2A/2B and related exclusion codes

**Criteria for Aim 4B Injury free, OA free controls are the same as in Aim 4A**.

###### Inclusions

- Probable idiopathic knee OA at any time (see Appendix 3B, i.e. ‘category 1’ knee OA from 3A, removing all post-traumatic knee OA codes) (NB when reporting this group’s characteristics, the first eligible OA code for this should be used)

###### Exclusions

- Evidence of an acute knee injury at any time (i.e. exclude all definite acute knee injury, defined by cohort as supported by Appendix 1A and related inclusion codes)
- A PTOA knee code at any time (Appendix 1B)
- Other simplified exclusions, defined by cohort, as supported by Appendix 2B

###### Criteria for Aim 4C Injury free, OA free controls are the same as in Aim 4A and 4B

Please populate the GO-PTOA Google sheet https://docs.google.com/spreadsheets/d/1JVrLm2dcsXRm_feyfixy7J621lkZJJz6VzNgix_JoyU/edit#gid=1072245856 with: 1) N for each group; 2) locally applied codes for inclusion and; 3) exclusion codes for each group. *Please complete table below if possible, noting if this analysis is not being carried out yet*.

Contact coordinator for final checks ahead of starting GWAS.

Upload your GWAS for:

- Aim 4A to folder **‘Aim4a_ALLPTOA_vs_HCONT’** on Box. Please include ‘Aim4a_ALLPTOA_vs_HCONT’ in the name of your file uploaded.
- Aim 4B to folder **‘Aim4b_KNEEINJ_vs_HCONT’** on Box. Please include ‘Aim4b_KNEEINJ_vs_HCONT’ in the name of your file uploaded.
- Aim 4C to folder **‘Aim4c_KIOA_vs_HCONT’** on Box. Please include ‘Aim4c_KIOA_vs_HCONT’ in the name of your file uploaded.

**Table 4.**
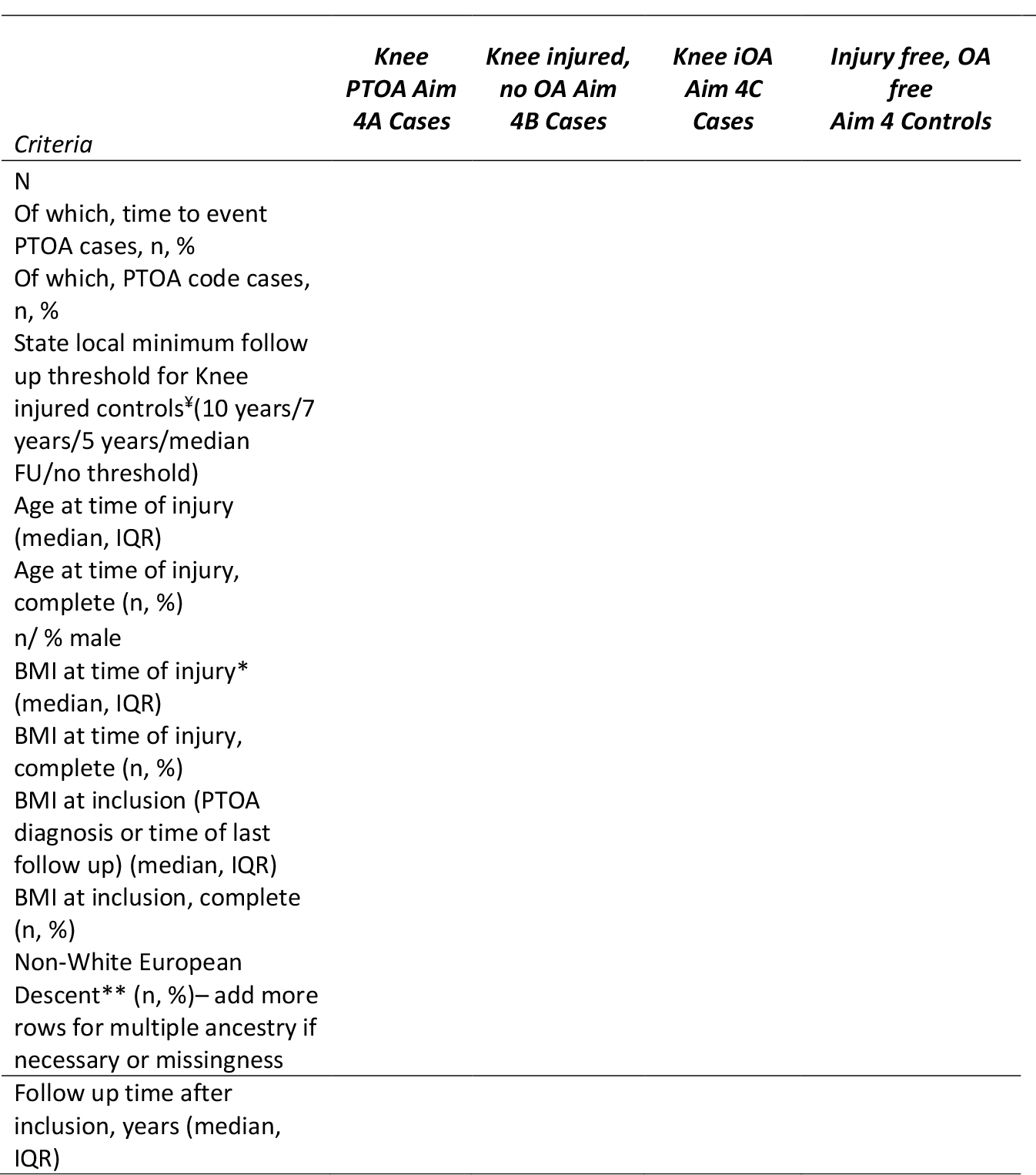
Aim 4 Descriptives. * BMI at time of injury, PTOA diagnosis or inclusion should be reported (including completeness) and used in all BMI sensitivity analyses **(section 5)**. ** If significant number non-White European descent subjects please specify breakdown by major ancestry. State NA if not known.

###### 4.0. Sign off to proceed

For each analysis, complete the associated table(s) for relevant analyses for your cohort <u>in this document</u> and upload to folder on Box and also populate GO-PTOA Google sheet relevant tabs with:

- Number of cases: N for each group following applying inclusions and exclusions
- Codes used: Locally applied codes for <u>inclusion and exclusion</u> for each group for that analysis
- Additional information: Intended software, file format, list of covariates used, number of genetic PCs & reference haplotypes for pre-imputation QC

Then contact group coordinator for final checks by coordinating and analysis teams ahead of starting GWAS (attaching this completed document or referring to Box upload as preferred).

###### 5.0. Covariates and methods for local analysis (see section 8.0)

Please be prepared to include listed covariates.

###### 6.0. Planned sensitivity analyses for all Aims

Where resources allow and this is possible, for each Aim please also run your model adjusting for BMI.

The group recognise that BMI could be on the causal pathway for osteoarthritis following injury and, its inclusion could lead to collider bias. The timing of collection of BMI may vary between cohorts. However, the group also feel that the effects of BMI are important to consider and likely to be a reviewer comment. As such we plan to investigate its effects on our findings in this planned sensitivity analysis. If its inclusion affects our findings, we will attempt to understand why and report this.

###### Aim 1 Identify genetic variation associated with knee PTOA independent of knee iOA

To achieve this aim, each cohort will perform a case-control GWAS of knee PTOA vs. knee iOA cases. *For the BMI sensitivity analyses, also perform analyses adjusting for BMI at time of OA diagnosis in study*.

Cohorts should upload their BMI-conditioned GWAS for Aim 1 to folder

**‘Aim1_ALLPTOA_vs_KIOA’** on Box. Cohorts should name this file

‘Aim1_ALLPTOA_vs_KIOA_BMI’.

###### Aim 2 Identify genetic variation significantly predictive of the onset of knee PTOA

Each cohort with available data will perform a time-to-event GWAS of all knee injured cases with available time information, looking at the subsequent event of presence of knee OA/PTOA, to 20 years of follow up or censor.

*For the BMI sensitivity analyses, please include all available BMIs for the subject during the 20 years of follow up*.

Cohorts should upload their BMI-conditioned GWAS for Aim 2 to folder

**‘Aim2_TEPTOA_vs_KNEEINJ’** on Box. Cohorts should name this file ‘Aim2_ TEPTOA_vs_KNEEINJ_BMI’.

###### Aim 3 Identify genetic variation associated with the onset of knee PTOA compared with knee injury controls without OA

Each cohort with available data will perform a case-control GWAS of knee PTOA cases vs. knee injured controls (no OA).

i. *For the BMI sensitivity analyses, please adjust for BMI at time of injury or if not available, at time of inclusion (last follow up for controls or OA diagnosis for cases)*.
ii. *For PTOA ascertainment sensitivity analyses, please exclude PTOA code cases that lack any prior injury information, or controls that have no documented follow-up time*.

Cohorts should upload their BMI-conditioned GWAS for Aim 3 to folder **‘Aim3_ALLPTOA_vs_KNEEINJ’** on Box. Cohorts should name this file ‘Aim3_ALLPTOA_vs_KNEEINJ_BMI’.

###### Aim 4 Identify genetic variation associated with knee PTOA compared with age matched controls without injury or OA

Each cohort with available data will perform :

A. case-control GWAS of knee PTOA cases versus injury and OA free controls
B. case-control GWAS of knee injured versus injury and OA free controls and
C. case-control GWAS of knee iOA vs. Injury free, OA free controls

Results from the three models will be compared to identify genetic variants likely to be associated with knee PTOA rather than knee injury risk or knee iOA.

*For all three models for the BMI sensitivity analyses, please use BMI at inclusion to be consistent across groups*.

Cohorts should upload their BMI-conditioned GWAS for:

- Aim 4A to folder **‘Aim4a_ALLPTOA_vs_HCONT’** on Box. Cohorts should name this file ‘Aim4a_ALLPTOA_vs_HCONT_BMI’
- Aim 4B to folder **‘Aim4b_KNEEINJ_vs_HCONT’** on Box. Cohorts should name this file ‘Aim4b_KNEEINJ_vs_HCONT_BMI’
- Aim 4C to folder **‘Aim4c_KIOA_vs_HCONT’** on Box. Cohorts should name this file ‘Aim4c_KIOA_vs_HCONT_BMI’

###### 7.0. Overarching principles of analysis execution and sharing of summary statistics

Each case-control analysis will be carried out in two stages, local analysis then upload and meta-analysis by the central site.

1. Each cohort will run their own individual GWAS for each aim where data and resources are available.
2. For the larger cohorts (especially when there is a case control imbalance) and resources permit, we recommend case-control GWAS analyses be performed using BOLT-LMM or SAIGE. For smaller cohorts, case-control analyses should be performed using tool team is most familiar with e.g. PLINK2, REGENIE This should be recorded in Google sheet
3. Recommended covariates to be used in case-control GWAS models: age, sex and local PCs for ancestry as appropriate. For BMI sensitivity analyses, either BMI at time of injury or BMI at PTOA/ OA diagnosis will also be used as a covariate.
4. Each cohort will share the genome-wide summary statistics for their analyses with the central analysis site, the University of Alabama at Birmingham (UAB), by uploading data to Box directory for their cohort/ study and Aim.
5. The Central analysis site (UAB) will carry out a single meta-analysis for each GWAS case control analysis.

###### Genome-wide array data

- Quality control the genotype data with the usual QC steps
- Account for relatedness is related individuals are in dataset at the analysis stage

###### Autosomal imputation

If you have your own reference haplotypes, then skip this section. If not, then we recommend the TOPMed panel. If you use an alternative, please just list and briefly explain this (please add to the Google Sheet). The current state of the art for imputation is to perform it using the BioDataCatalyst TOPMed imputation server which houses the TOPMed reference panel.

https://imputation.biodatacatalyst.nhlbi.nih.gov/#!pages/home

Comprehensive details regarding imputation using the BioDataCatalyst imputation server are covered here: https://topmedimpute.readthedocs.io/en/latest/

###### Combine datasets if necessary

In situations where cases and controls are genotypes separately, the variants should be aligned <u>to GRCh 38 and forward strand</u>.

The cases and controls should be combined into a single dataset to include only the overlapping variants. Additional QC will be prudent following this, for example: Generate and visualize PCs, check for duplicates, differential missingness, inspection of intensity clusters for significant variants after case-control association analysis.

###### Pre-imputation QC

Pre-imputation QC can be assessed using BioDataCatalyst imputation server and selecting quality control only under ‘Mode’ which will generate a QC report for your review prior to initiating imputation. However, we recommend additional pre-imputation QC exclusing AT/GC variants as well as those deviating from Hardy Weinburg equilibrium P<1E-4. This QC should be performed prior to uploading to the imputation server which has its own QC pipeline where each variant is checked and excluded if:

1. contains invalid alleles
2. duplicates
3. indels
4. monomorphic sites
5. allele mismatch between reference panel and uploaded data
6. SNP call rate <90%

The filtered variants are output in a file called statistics.txt for your review. Additional details on formatting file for upload to the imputation servers are here: https://topmedimpute.readthedocs.io/en/latest/prepare-your-data.html

###### Post-imputation QC

There is no need to exclude variants based on Hardy-Weinberg Equilibrium p-value or imputation information score as this can be performed centrally.

###### Autosomal association analysis

If your cohort contains individuals from different ancestries, please can you analyze these ethnicities separately. Please email if you have questions regarding ethnic groupings and/or sample sizes.

###### Software

The following software can be used:

- BOLT-LMM (https://alkesgroup.broadinstitute.org/BOLT-LMM/)
- REGENIE (https://rgcgithub.github.io/regenie/)
- SAIGE (https://github.com/weizhouUMICH/SAIGE)
- GEMMA (https://github.com/genetics-statistics/GEMMA)
- RVTest (http://zhanxw.github.io/rvtests/)
- SNPTEST (https://mathgen.stats.ox.ac.uk/genetics_software/snptest/snptest.html)

If you need to use a mixed model, are limited computationally or have large cohorts then BOLT-LMM is better than SNPTEST. If using SNPTEST please apply—method score

###### Covariates

Please include age (at time of injury or age at time of inclusion as appropriate) and sex, Genetics PCs and any cohort covariates as appropriate. **Only include BMI as a covariate for sensitivity analyses**.

- Age at time of injury
- Or, Age at time of inclusion (diagnosis/last follow up – see analysis)
- Sex
- Global ancestry group (if different ancestral groups are included)
- First X genetic principal components (as defined by cohort)
- Any other cohort specific covariates, felt to be important by cohort, such as Genotyping chip

Update the Additional Information tab of the Google sheet on the covariates you use. https://docs.google.com/spreadsheets/d/1JVrLm2dcsXRm_feyfixy7J621lkZJJz6VzNgix_JoyU/edit#gid=0

###### Autosomal data upload

Please upload genome-wide summary statistics as a <u>tab delimited text file</u> labelled with the following format (See Table 1 for abbreviations): **STUDY_PHENO_ETHNICITY_NCASES_NCONTROLS_SOFTWARE_PANEL_BUILD_DATE_ANALYST_GC_FF.txt**

(E.g. Aim1_ALLPTOA_vs_KIOA_EA_8078CASES_9268CONTROLS_SNPTEST_TOPMED_38_070523_LOZ_GCNo_FFNo.txt)

If you are uploading imputation information scores separately please prefix with INFO as so:

**INFO_STUDY_PHENO_ETHNICITY_NCASES_NCONTROLS_SOFTWARE_PANEL_BUILD_DATE_ANALYST_GC_FF.txt**

(E.g. INFO_Aim1_ALLPTOA_vs_KIOA_EA_8078CASES_9268CONTROLS_SNPTEST_TOPMED_38_070523 _LOZ_GCNo_FFNo.txt)

If you are uploading imputation information scores separately please prefix with INFO as so:

**INFO_STUDY_PHENO_ETHNICITY_NCASES_NCONTROLS_SOFTWARE_PANEL_BUILD_DATE_ANALYST_GC_FF.txt**

(E.g.INFO_ALLPTOA_vs_KIOA_EA_8078CASES_9268CONTROLS_SNPTEST_TOPMED_38_070523 _LOZ_GCNo_FFNo.txt)

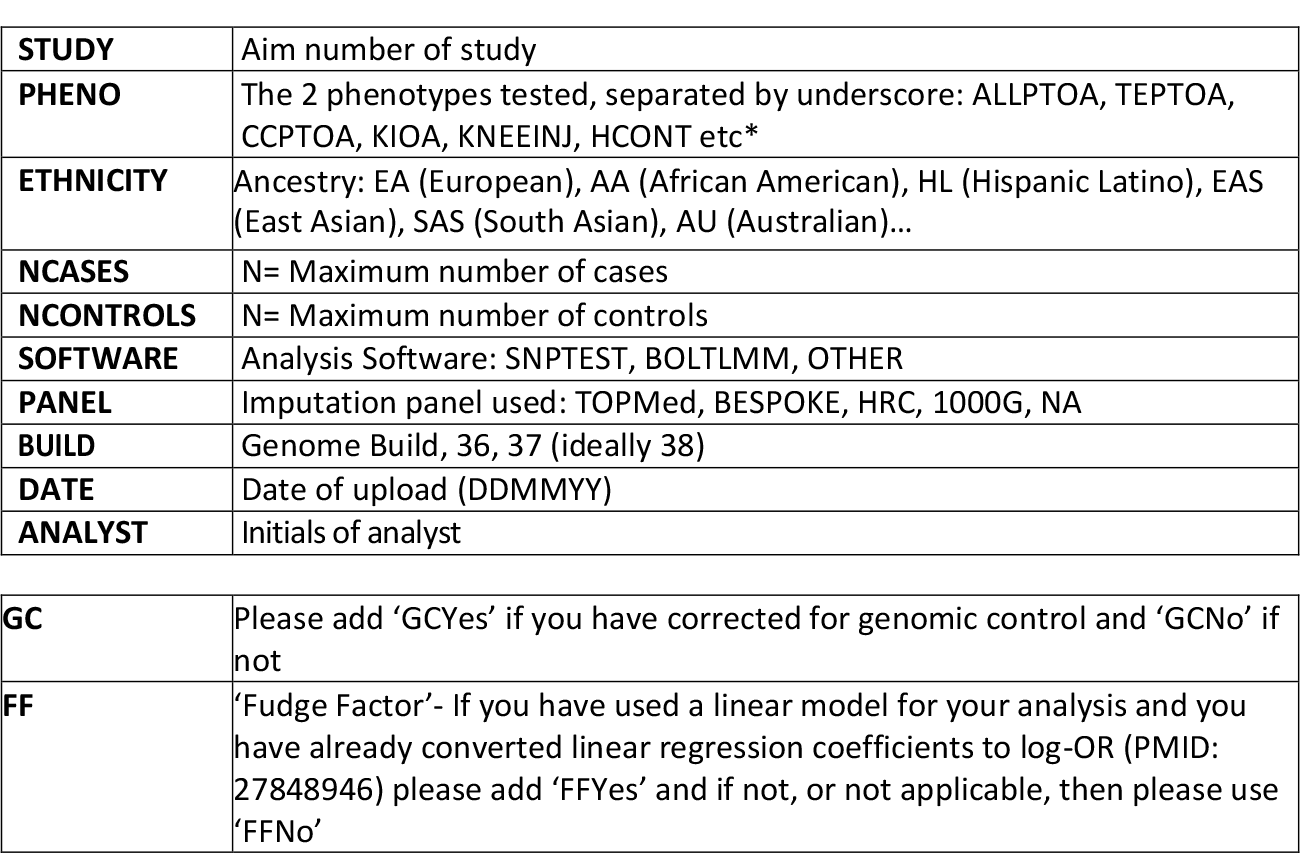

###### Requested summary statistics

Please upload the following association analysis summary statistics. If you have BOLT-LMM or SNPTEST then uploading the output files directly is fine. Imputation information score can be uploaded separately but please include your unique marker identifier along with this.

If you have used alternative software or have performed any additional reformatting, please upload the following. Please note that the column naming is important but the column order is not important and uploading additional columns will be fine.

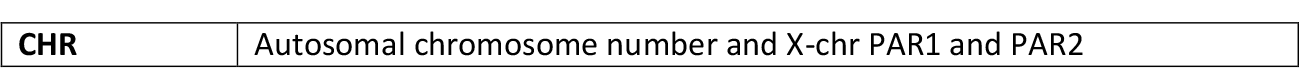
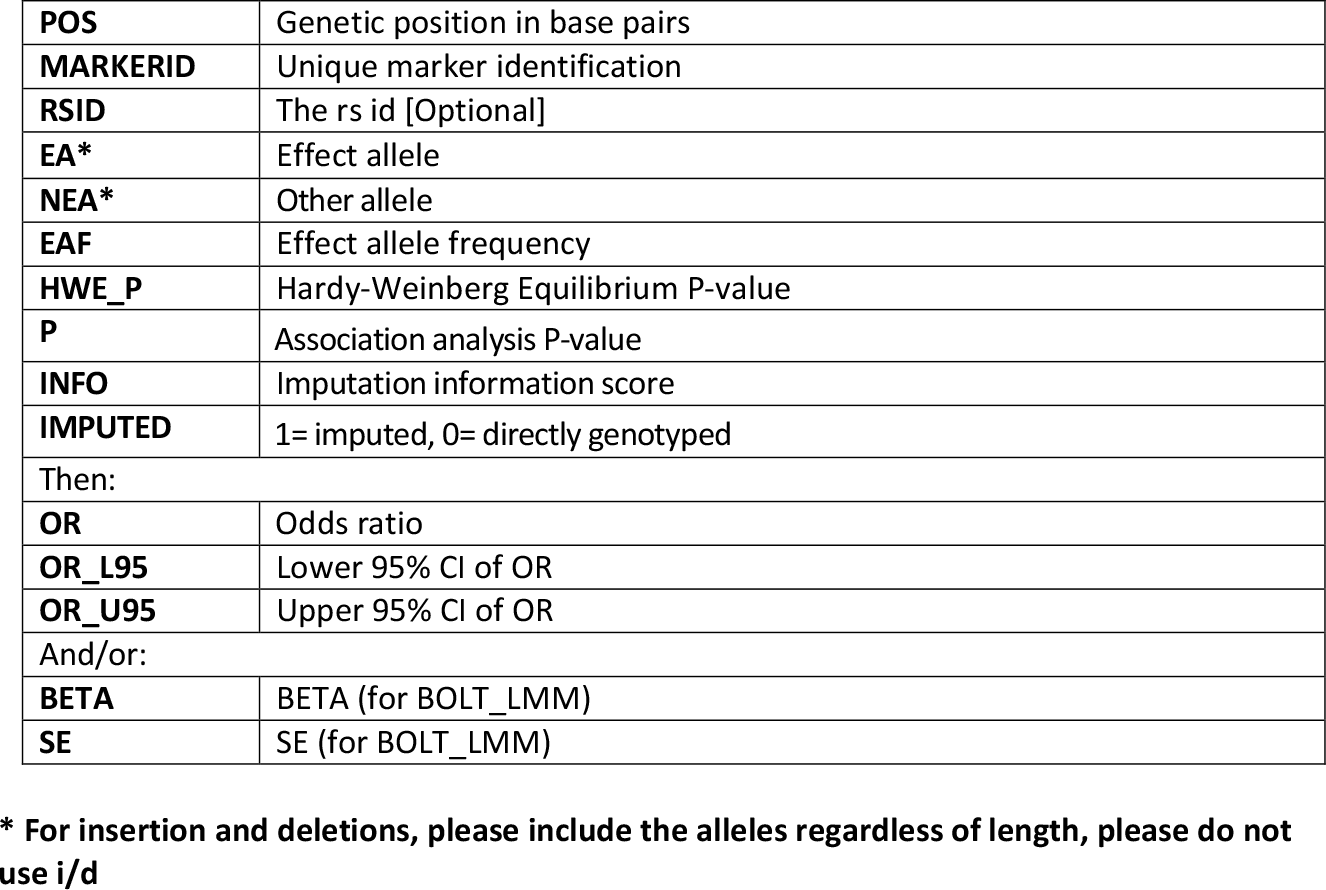

###### Upload to Box

We will use provided email addresses for all analysts to set up access to your cohort/study Box folder(s) which will each contain a folder for each analysis. UAB will issue a Box link accordingly. Please include a file containing the md5sum results for each file uploaded using the following example as a guide: **md5sum file1.txt file2.txt file3.txt > STUDY.md5.txt**

###### Pre-imputation genotyping quality control (QC): for the directly typed variants

In situations where cases and controls are genotyped separately, the variants should be aligned to the appropriate build and forward strand. The cases and controls should be combined into a single dataset to include only the overlapping variants. Additional QC will be prudent following this, for example: Check for duplicates, differential missingness, inspection of intensity clusters for significant variants after case-control association analysis.

In addition to QC outlined below, if you are newly performing the X chromosome imputation, we highly recommend the use of Michigan Imputation Server pre-QC and imputation which houses all relevant reference panels other than your own if you need to use a private reference panel or TOPMed which is housed on the BioDataCatalyst TOPMed imputations server. The chromosome X pipeline performs additional QC on the X chromosome in addition to splitting the chromosome into three independent chunks (PAR1, nonPAR and PAR2).

https://imputationserver.sph.umich.edu/index.html#!pages/home

https://imputation.biodatacatalyst.nhlbi.nih.gov/#!pages/home

We are working with Build 38. These are the coordinates for the pseudo autosomal (PAR1 and PAR2) and non-pseudo autosomal (nonPAR) regions on X chromosome.

| Chr X region | Coordinates in bp (b38) |
| --- | --- |
| PAR1 | chr X1 (< 2781479) |
| nonPAR | chr X2 (2781479 - 155701383) |
| PAR2 | chr X3 (> 155701383) |

Split the X-chromosome directly typed data into 2 datasets:

1. **PAR1 and PAR2**
2. **nonPAR**

We will QC these separately, as detailed below and then we combine at the end of this section. Example: In plink the data Xchr data can be extracted and separated into nonPAR and PAR12 by the following (here your genome-wide dataset = data.bed/bim/fam). Note that there may be no variants in the PAR12 region in your dataset.

plinkv1.9 --bfile data --chr 23 --split-x b38 no-fail --make-bed --out chrX

###### (1) PAR1 and PAR2

Please QC exactly as you did for the autosomes. Example: In plink this can be achieved by the following (chrX data was produced as above) we use an exclusion threshold of HWE p<1×10^−4^, 1% MAF and 98% call rate.

plinkv1.9 --bfile chrX --chr 25 --hwe 0.0001 --make-bed --geno 0.98 --maf 0.01

--out QCedPAR12_data

###### (2) nonPAR

Please split your cohort into males and females for the QC:

- In the female dataset perform your Hardy-Weinberg p-value filters and create a variant inclusions list (inclusions1)

Example: In plink this can be achieved by the following (chrX data was produced as above) for an exclusion threshold of HWE p<1×10^−4^

plinkv1.9 --bfile chrX --filter-females --chr 23 --hwe 0.0001 --make-bed --out female_nonPAR_data

cut -f 2 female_nonPAR_data.bim > Inclusions1

- In the male dataset prepare a list of variants that contain individuals that are heterozygous haploids prepare an exclusions list for these (Inclusions2)

Example: In plink this can be achieved by the following:

plinkv1.9 --bfile chrX --filter-males --chr 23 --from-bp 2699520 --to-bp 154931044 --make-bed --out male_nonPAR_data

If you have no male_nonPAR_data.hh file produced then you have no heterozygous haploids

in your data and there are no exclusions to prepare. But if you do…

cut -f 3 male_nonPAR_data.hh | sort -u > Exclusions1

plinkv1.9 --bfile male_nonPAR_data --exclude Exclusions1 –make-bed –out male_nonPAR_data2

cut -f 2 male_nonPAR_data2.bim > Inclusions2

Include only variants that pass both QC in males and females (Inclusion3).

Example: to get the intersect from both lists you can use the following:

comm -1 -2 <(sort Inclusions1) <(sort Inclusions2) > Inclusions3

Extract these variants from the chrX file and include your usual call rate and MAF filters. Example: In plink this can be achieved by the following, here with a 1% MAF and 98% call rate threshold:

plinkv1.9 --bfile chrX --extract Inclusions3 --geno 0.98 --maf 0.01 --make-bed

--out QCednonPAR_data

###### (3) Combine the QCed PAR1, PAR2, nonPAR into a single file and code chrX as 23

Males and females should be in the same file and the Xchr should be coded as 23 regardless if the variants are in the nonPAR or PAR region. This is because the imputation server need everything as 23 or X. example: In plink this can be achieved by the following, with the QCed data prepared and files name as above.

plinkv1.9 --bfile QCednonPAR_data --bmerge QCedPAR12_data --make-bed -- out QCedXchr_data

plinkv1.9 --bfile QCedXchr_data --merge-x no-fail --make-bed --out QCedXchr_data

Quality control for HRC or older reference panels requires comparison of alleles and frequencies to reference panel. Details are here: https://topmedimpute.readthedocs.io/en/latest/prepare-your-data.html

For TOPMed imputation, comparison to the TOPMed reference panel is performed within the imputation pre-QC. Details are here: https://topmedimpute.readthedocs.io/en/latest/pipeline.html

You will then need to prepare vcf files for upload as detailed in the link above.

###### Imputation

Please make a note of the number of directly types variants as we will ask that you include this information in your data upload. Example: imputation with HRC haplotypes on the Michigan server for a European cohort.

|  |  |
| --- | --- |
| Reference panel: | TOPMed r2 (hg38) |
| Array Build: | GRCH38/hg38 |
| Rsq filter: | off |
| Phasing: | Eagle v2.4 (phased output) |
| Population:: | EUR |
| Mode: | Quality Control & Imputation |

###### Post imputation QC: for the imputed variants

We are stringent regarding the variants we include for chrX analysis. To summarise the process:

- Split chrX into nonPAR and PAR1+2
- For the nonPAR, we exclude the following:
  - Genotypes with genotype probability <0.9
  - Variants with call rate <99%
  - Variants with ≥ 1 heterozygote haploid genotype in males
  - Variants with Hardy-Weinberg p<0.0001 in females

###### PAR1+2

For the PAR1 and 2 regions we extract the variants located in these regions but filter out variants with an estimated imputation accuracy (R-square) < 0.3. This data is ready to use for association analysis using the same protocol you applied for the autosomal analysis.

Example: Where the imputed vcf from HRC imputation server is named chrX.dose.vcf.gx.

tabix -f -p vcf chrX.dose.vcf.gz

bcftools view -r X:1-2699519,X:154931045-15527056 ‘-e INFO/R2<0.3’ -Oz –o chrX.PAR12.filteredR2less0.3.vcf.gz

chrX.dose.vcf.gz tabix -f -p vcf

chrX.PAR12.filteredR2less0.3.vcf.gz

###### nonPAR

For the nonPAR regions we apply very stringent QC, we extract all variants in the region with R2 ≥ 0.3 and then we will apply additional QC. Example:

bcftools view -r X:2699520-154931044 ‘-e INFO/R2<0.3’ -Oz -o chrX.nonPAR.filteredR2less0.3.vcf.gz chrX.dose.vcf.gz

tabix -f -p vcf chrX.nonPAR.filteredR2less0.3.vcf.gz

To ensure that the fam file matches the vcf you may have to prepare double identified for Plink. Example: where the fam file produced as the end of step 2 above (*) is called QCedXchr_data-updated-chr23.fam

cat QCedXchr_data-updated-chr23.fam | awk ‘{print

$1”_”$1,$2”_”$2,$3,$4,$5,$6;}’ > chrX.dose.fam

Exclude any genotypes that do not have a high genotype probability. Convert the genotype probabilities into hard calls using a genotype probability threshold of 0.9. Example: Note that we switch to plink v2.0 for this step.

plinkv2 --vcf chrX.nonPAR.filteredR2less0.3.vcf.gz dosage=GP --double-id

-- import-dosage-certainty 0.9 --keep-females --make-bed –out

female_chrX.nonPAR_GP0.9 --psam chrX.dose.fam

plinkv2 --vcf chrX.nonPAR.filteredR2less0.3.vcf.gz dosage=GP --double-id

-- import-dosage-certainty 0.9 --keep-males --make-bed --out

male_chrX.nonPAR_GP0.9 --psam chrX.dose.fam

Apply a call rate threshold of 99% and HWE p<1 × 10^−4^ for the females. Example:

plinkv1.9 --bfile female_chrX.nonPAR_GP0.9 --geno 0.01 --hwe 0.0001 --make-bed

--out female_chrX.nonPAR_GP0.9_geno0.01_pHWE1e-4

Apply a call rate threshold of 99% for the males. Example:

plinkv1.9 --bfile male_chrX.nonPAR_GP0.9 --geno 0.01 --make-bed --out

male_chrX.nonPAR_GP0.9_geno0.01

If you do not have a .hh file produced then skip the next 2 example steps and for the males use the mail bim file produced in the step above for the inclusions.

If you have a .hh file produced then there are variants with incorrect genotypes (heterozygous haploids) so we will exclude all of these variants. Example:

cat male_chrX.nonPAR_GP0.9_geno0.01.hh | cut -f 3 | sort -u >

male_chrX.nonPAR_GP0.9_geno0.01.hhexclusions

Exclude the variants with heterozygous haploids. Examples:

plinkv1.9 --bfile male_chrX.nonPAR_GP0.9_geno0.01 –exclude

male_chrX.nonPAR_GP0.9_geno0.01.hhexclusions --make-bed --out

male_chrX.nonPAR_GP0.9_geno0.01_hhout

Produce an inclusions list for the variants that pass QC in males and females. Example: to get the intersect from both bim files you can use the following:

comm -1 -2 <(cut -f 2 male_chrX.nonPAR_GP0.9_geno0.01_hhout.bim | sort)

<(cut - f 2 female_chrX.nonPAR_GP0.9_geno0.01_pHWE1e-4.bim | sort) > nonPARQCedinclusions.txt

Prepare sex specific datasets for association analysis

plinkv1.9 --bfile male_chrX.nonPAR_GP0.9_geno0.01 –extract

nonPARQCedinclusions.txt --make-bed --out male_chrXnonPAR_QCed

plinkv1.9 --bfile female_chrX.nonPAR_GP0.9_geno0.01_pHWE1e-4 –extract

nonPARQCedinclusions.txt --make-bed --out female_chrXnonPAR_QCed

Because we have set some genotypes to missing it is good practice to make sure that you do not have differentially missing genotypes between your cases and controls for each phenotype.

Example for phenotype 1, which is included in column 6 of the fam file:

plinkv1.9 --bfile female_chrXnonPAR_QCed

--test-missing --out female_chrXnonPAR_QCed_diffMiss_pheno1

Exclude variants with P<1×10^−4^

cat female_chrXnonPAR_QCed_diffMiss_pheno1.missing | awk ‘{if($5 <0.0001) print

$2;}’ > female_chrXnonPAR_QCed_diffMiss_pheno1

These variants should be excluded from your association analysis. Example:

plink --bfile female_chrXnonPAR_QCed –-exclude

female_chrXnonPAR_QCed_diffMiss_pheno1 -–make-bed –-out final_file_pheno1

This means that if you have >1 phenotype the variants excluded may be different for each phenotype.

Using plinkv2.0 with --export you can also prepare these genotypes as vcf, bgen or Oxford format to facilitate your association analysis requirements.

###### X-chromosome association analysis

The X chromosome will be analysed as 3 separate datasets:

- PAR1+2
  - Analyse as you did for the autosomal analysis and follow the autosomal analysis plan for file naming and uploading.
- nonPAR males only
- nonPAR females only
  - Analyse as you did for the autosomal analysis but follow the file naming and uploading instructions below

Please note that centrally, we will:

- Investigate if we need to correct for genomic control
- Apply quality control
- Meta-analyse the male and female data centrally using Z-scores

###### X-chromosome data upload

If you performed imputation with HRC or 1000Genomes please can you also upload the html file from step 4 above, using the following naming convention **STUDY_ETHNICITY_PANEL_BUILD_NSCAFFOLD_DATE_ANALYST.html**

Please upload nonPAR summary statistics as a tab delimited text file labelled with the following format. Note that there are differences to the autosomal analysis naming protocol (highlighted in red): **STUDY_SEX_PHENO_PHENO_ETHNICITY_NCASES_NCONTROLS_SOFTWARE_PANEL_BUILD_NSCAFFOLD_REL_DATE_ANALYST_GC_FF.txt**

(E.g.OASTUDY_FEMALEPTOA_EA_407CASES_5268CONTROLS_SNPTEST_TOPMED_38_1231_ UNREL_070218_LOZ_GCNo_FFNo.txt)

|  |  |
| --- | --- |
| **STUDY** | Aim of study |
| **SEX** | Is it the male only or female only analysis: MALE, FEMALE |
| **PHENO** | The phenotypes compared separated by underscore: ALLPTOA, TEPTOA, CCPTOA, KIOA, KNEEINJ, HCONT etc. |
| **ETHNICITY** | Ancestry: EA (European), AA (African), HL (Hispanic Latino), EAS (East Asian), SAS (South Asian), AU (Australian) |
| **NCASES** | N= Maximum number of cases |
| **NCONTROLS** | N= Maximum number of controls |
| **SOFTWARE** | Analysis software: SNPTEST, BOLTMM, OTHER |
| **PANEL** | Imputation panel used: BESPOKE, HRC, 1000G, NA |
| **BUILD** | Genome build: 36, 37, 38 (Note that we are using 38) |
| **NSCAFFOLD** | The number of directly types chrX variants used for the imputation |
| **REL** | Are there related (RED) individuals in your cohort or not (UNREL): REL UNREL, we will account for the related samples between the males and females in your cohort centrally. |
| **DATE** | Date of upload (DDMMYY) |
| **ANALYST** | Initials of analyst |
| **GC** | We will test and correct for genomic control centrally. Therefore please add 'GCNo'. If you need to apply this yourself add 'GCYes' |
| **FF** | 'Fudge Factor'- If you have used a linear model for your analysis and you have already converted linear regression coefficients to log-OR (PMID: 27848946) please add 'FFYes' and if not, or not applicable, then please use 'FFNo' |

###### Requested summary statistics

Please upload to your Box folder the following association analysis summary statistics. Please note that the column naming is important but the column order is not and uploading additional columns will be fine.

|  |  |
| --- | --- |
| **CHR** | Chromosome number |
| **POS** | Genetic position in base pairs |
| **MARKERID** | Unique marker identification |
| **RSID** | The rs id [Optional] |
| **EA\*** | Effect allele |
| **NEA\*** | Other allele |
| **EAF** | Effect allele frequency |
| **HWE_P** | Hardy-Weinberg Equilibrium P-value |
| **NCASES** | The number of cases |
| **NCONTROLS** | The number of controls |
| **P** | Association analysis P-value |
| **INFO** | Imputation information score |
| **IMPUTED** | 1= imputed, 0= directly genotyped |
| Then: |  |
| **OR** | Odds ratio |
| **OR_L95** | Lower 95% CI of OR |
| **OR_U95** | Upper 95% CI of OR |
| And/or: |  |
| **BETA** | Beta (for BOLT_LMM) |
| **SE** | SE (for BOLT_LMM) |

\* For insertion and deletions please include the alleles regardless of length and please do not use i/d

Please include a file containing the md5sum results for each file uploaded using the following example as a guide: md5sum file1.

This Appendix 5.0 relates to the GO-PTOA analysis plan, Version 2.0. Please refer to the main document for all other queries.

Summary of amendments to V1.0 of the Analysis Plan (dated 28^th^ April 2023), resulting in V2.0 (dated 18^th^ August 2023):

1. Addition of contents page at the start of the Analysis Plan
2. Addition of references to code list documents to Appendices 1, 2 & 3 (footnotes)
3. Clarification on summary statistics file naming convention for upload throughout section 3.0 of Appendix 5, including file naming for both phenotypes and for BMI-conditioned GWAS for sensitivity analyses in section 6.0
4. Updated examples of file names in the Autosomal data upload segment of section 8.0 of Appendix 5

## Footnotes

1 Refer to Excel/ Text files ‘ICD9 Inclusions (Appendix 1)_V1.0_27 May 23’ and ‘ICD10 Inclusions (Appendix 1)_V1.0_27 May 23’

2 Refer to Excel/Text files ‘ICD10 Exclusions (Appendix 2A & 2B)_V1.0_27 May 23’

3 Refer to Excel/Text files ‘ICD10 Exclusions (Appendix 2A & 2B)_V1.0_27 May 23’ & Excel/Text files ‘ICD9 Exclusions (Appendix 2B)_V1.0_27 May 23’

4 Refer to Text files ‘ICD9 3A ALLKOA_V1.0_9 June 23’ & ‘ICD10 3A ALLKOA_V1.0_9 June 23’ and Excel File ‘ICD10 Cat 1 osteoarthritis outcome codes_(Appendix 3)_V1.0_9 June 23’

5 Refer to Text files ‘ICD9 3B KIOA_V1.0_27 May 23’ & ‘ICD10 3B KIOA_V1.0_27 May 23’

